# Transcription factor binding and individual genetic risk for valproate teratogenicity

**DOI:** 10.1101/2025.02.24.25322812

**Authors:** Alison Anderson, Piero Perucca, Elena Vianca, Danial Sandvik, Ana Antonic-Baker, Roland Krause, Dana Jazayeri, Alison Hitchcock, Janet Graham, Marian Todaro, Torbjörn Tomson, Dina Battino, Emilio Perucca, Meritxell Martinez Ferri, Anne Rochtus, Lieven Lagae, Maria Paola Canevini, Elena Zambrelli, Ellen Campbell, Aleksei Rakitin, Bobby P. C. Koeleman, Ingrid E. Scheffer, Samuel F. Berkovic, Patrick Kwan, Sanjay M. Sisodiya, John Craig, Frank J. E. Vajda, Terence J. O’Brien, the EpiPGX and EPIGEN Consortia

## Abstract

Valproate (VPA) use during pregnancy is associated with a wide range of birth defects and adverse neurodevelopmental outcomes, but not all exposed children are affected and there is evidence for a genetic predisposition. We hypothesised that genomic variants that impact on the binding affinity of transcription factors (TFs) are integral to VPA-associated teratogenicity and a plausible explanation for variance in interindividual risk. We interrogated maternal exomes from women recruited through international epilepsy genomics consortia. The variant burden within genes associated with 32 different birth defect types was higher for those exposed to VPA as compared to other antiseizure medications (OR 1·73 [95% CI 1·40 to 2·14], *p* = 2·25E-07). Variants in women exposed to VPA were predicted to impact the binding affinity of 359 TFs and network analysis of encoded proteins indicated that a master regulator, *EP300,* interacts with 42% (151/359) of all variant sensitive TFs. We then profiled coexpression between *EP300* and other TFs in differentiating neurons derived from human embryonic stem cells (hESCs) exposed to VPA at 300µM and 700 µM, or unexposed, and a reference map generated using public data. We found strong overlap in *EP300*-TF coexpressed pairs between the reference and all comparison groups (99%,900/911) but only 32% (134/422) of pairs observed in unexposed cells were evident following VPA exposure, and over half of all pairs (489/911) were observed in VPA-exposed cells only. Our findings suggest that VPA-induced disruption of *EP300-*related regulation is common across birth defect types and that genetic variation can modify subsequent transcriptional dysregulation, explaining why only some pregnancies are affected. The results have implications for the development of genetic risk biomarkers and safer drugs.

## Introduction

Valproic acid (VPA) is a first-line anti-seizure medication (ASM) for the control of seizures in people with epilepsy, in particular those with genetic generalised epilepsies where it is the most effective treatment.^1, 2^ It is also widely used for the treatment of neuropathic pain, bipolar disorder, schizophrenia, migraine^3^ and is emerging as a potential cancer therapy.^4^ Babies born to mothers who take VPA during pregnancy are at high risk of birth defects, lower intelligence quotient (IQ), developmental delay, attention deficit hyperactivity disorder, autism spectrum disorder, and other neurodevelopmental disorders.^3^ Globally, measures have been introduced to raise awareness and restrict VPA prescribing in females of childbearing potential, and in some countries this restriction has been extended to males.^5^ Yet, not all children exposed *in utero* to VPA have adverse consequences, and particularly for patients with generalised epilepsies, alternatives to this potentially life-saving medication are often considerably less effective.^1, 2^ The teratogenic effects have been well-established through large prospective registries^6, 7^, administrative healthcare databases,^8, 9, 10^ and prospective, smaller-scale, observational studies.^11, 12^ There remains a pressing need to elucidate the mechanism of teratogenesis and to understand risk at the molecular and individual level. A genetic susceptibility to VPA teratogenicity is suggested by studies that found increased risk of recurrent ASM-associated birth defects in women who have had an affected pregnancy,^13^ those with a family history of birth defects,^14^ and animal models.^15^

The nature of VPA teratogenicity is highly variable and includes non-specific structural birth defects affecting multiple different organ systems, as well as neurodevelopmental deficits.^16^ During organogenesis and embryonic development, the expression of genes is tightly orchestrated in time and space by DNA-binding proteins collectively referred to as transcription factors (TF).^17^ The binding of TFs to DNA is dependent on both specific amino acid residues in the TF protein and specific bases in the target DNA sequence. Genetic variants within TF binding sites can (i) modify binding affinity strength, (ii) prevent one or more TFs from binding, or (ii) introduce a motif that permits aberrant TF binding.^18^

Genome-wide association studies (GWAS) show that polymorphisms in TF binding sites comprise only 8% of the genome, yet represent 31% of all trait-associated polymorphisms.^19^ These variants are mostly located within intronic (∼50%) and intergenic (∼30%) DNA regions, but are also found in promoters, coding regions and untranslated gene regions^18^. It has been shown that enhancers (regulatory DNA regions) with tissue-specific activity are enriched in intronic regions^20^. High quality genotypes within intronic and intergenic variants are captured by the approximately 40-60% of exome-sequencing reads that are off-target.^21^ Here, we used exome data to explore the hypothesis that variant-induced changes in TF binding might modify VPA-induced dysregulation of gene expression and thus explain both differences in interindividual risk and the heterogeneity in VPA-associated birth defects. We analysed and compared pregnancies exposed to VPA only and non-VPA ASMs, ascertained from epilepsy-pregnancy registers and applied a network-based approach that contextualised the variant spectra to genes associated with birth defects and integrated evidence from multiple modalities, to test our proposed mechanism of teratogenesis.

## Materials and Methods

### Samples

Women were recruited through an international collaboration involving centers in Australia, Europe and North America: The Raoul Wallenberg Australian Pregnancy Register of Antiepileptic Drugs,^22^ The UK and Ireland Epilepsy and Pregnancy Register (UKIEPR), Epilepsy Pharmacogenomics Consortium (EpiPGX), and an international epilepsy genetics research consortium, EPIGEN.^23^ Blood sample-derived whole-exome data were available for mothers treated with VPA, or other ASMs during pregnancy who had a termination due to birth defects, a livebirth with one or more congenital malformations, or a livebirth with no congenital malformations or identified neurodevelopmental problems. Exome sequencing was carried out through either the Broad Institute, the Institute for Genomic Medicine, Columbia University in New York, or deCODE genetics. The raw reads were aligned to human genome build GRCh38, and standard quality filtering and imputation methods applied [Supplementary methods]. To ensure that variant detection had not been biased by differences in the exome capture kits used, we compared (Wilcoxon signed-rank test) the median depth at the loci of high-confidence variants with 100 base-pair padding either side across capture kits.

### Association tests

The MalaCards human disease database was used to identify genes associated with ‘congenital structural birth defects’ or ‘congenital genitourinary birth defects’, which covered all of the birth defects of affected children reported in our study cohort. Gene burden association tests were undertaken using the SNP-Set (Sequence) Kernel Association Tests available in the ‘SKAT’ R package (version 2.2.4). These methods aggregate individual SNP score statistics in a SNP set, in this case each gene, and tests include Burden, SKAT and SKAT-O, which allows testing for common only, rare only, or common and rare variant associations. The tests have different strengths that depend on the underlying biology thus six different tests were applied and genes with *p* <0.05 in at least one test were included in subsequent analyses. Variant level association tests were undertaken using the Plink software version 2.0, with ten principal components to control for population stratification, and permutation to determine significance levels empirically. Fisher’s exact test was used to compare the proportion of significant genes, and unique gene-birth defect-type associations identified between sample subsets (VPA exposures or non-VPA ASM exposures).

### Network construction and knowledge integration

The STRING database App within the Cytoscape software (version v3.10.2) was used to generate protein-protein interaction networks from proteins encoded by significant genes, hereinafter referred to as the birth defect hub (BD-hub) for VPA exposures and non-VPA ASM exposures, and a network of TFs (TF-hub) that were predicted to have their binding affinity modified by one or more variants evidenced in the VPA exposure BD-hub. To infer the functional consequence of variants, we integrated evidence from: (i) the Ensembl variant effect predictor (VEP) tool, (ii) expression quantitative trait loci (eQTL) from the GTEX compendium, and (iii) evidence of potential disruption of TF binding sites using the DeepBind software tool (Supplementary methods, Fig. S3). To gain insight into the susceptibility of genes to VPA, we integrated findings from differential expression analyses as reported in a rodent model^24^ and a human cortical organoid model^25^ of VPA teratogenesis, and a human embryonic stem cell-based neurodevelopmental toxicity assay.^26^ In addition, we (i) used data from the neurodevelopmental toxicity assay to identify BD-hub genes that were differentially expressed by carbamazepine (CBZ) which can also have teratogenic effects, and (ii) assessed how many BD-hub genes overlapped a teratogenic ‘gene signature’ derived from a human induced pluripotent stem cell (hiPSC)-based assay for the classification of developmental toxicants.^27^

To establish the specificity of signal, we generated 1000 random gene sets comprising 44 genes (the median number of genes in gene sets associated with a specific birth defect). Using data from VPA-exposed pregnancies, we then compared the distribution of P-values, and the lowest P-value observed for each SKAT test between birth defect-related set genes and random set genes. Metrics representing the functional impact of variants in genes found to be significant across VPA-exposed pregnancies, non-VPA-exposed pregnancies, and 100 random gene sets were compared using a one sample t-test or Wilcoxon signed-rank test, depending on normality of the distribution.

### Variant prioritisation

Individual variants in genes within the BD-hub derived from VPA-exposed pregnancies were filtered based on evidence of functional consequence and considered to have a ‘high-level’ of evidence if all of the following criteria were met: (i) the variant was non-imputed, (ii) was independently associated with an adverse pregnancy outcome (Plink association test, empirical *p* ≤= 0·05), (iii) the eQTL effect for the variant included the gene that the variant lies within, (iv) DeepBind scores indicate that the variant impacts the binding of at least one TF, and (v) there was evidence from at least one exposure study that VPA modified the expression of the gene harbouring the variant, or one or more of the TFs predicted to be modified by the variant. Variant filtering was undertaken using R version R/4.2.0. Circos plots were constructed using the R circlize library.

### Reference map for *EP300* coexpression

To gain insight into how *EP300* interacts with other TFs we constructed a reference map using coexpression analyses and publicly available gene expression data. Processed bulk RNA-seq data from post-mortem brain tissue, representing the full course of human brain development, including both prenatal and postnatal periods, was downloaded from the BrainSpan Atlas of the Developing Human Brain (https://www.brainspan.org/). This resource provides a list of TFs which we supplemented with a list of human TFs, obtained from the Gene Transcription Regulation Database (GTRD), giving a total of 2,422 TFs for analyses. The correlation between *EP300* expression and the expression of all TFs from the list that were detected in the BrainSpan dataset was determined using both Pearson (to detect linear correlations in normally distributed data) and Spearman (to detect non-linear relationships) methods. *p* < 0.05 for either test was considered a true correlation.

In addition, single cell RNA-seq (scRNA-seq) data was used to profile coexpression in cells from Cerebellum and Cerebrum foetal tissue data obtained from the Gene Expression Omnibus repository (Accession number GSE156793). For this data type, the high dimensional Weighted Gene Coexpression Network Analysis (hdWGCNA) method was applied. This method overcomes the limitation of sparse single cell data by first constructing metacells (clusters of genes with similar expression identified using the K-Nearest-Neighbors (KNN) algorithm) and using the average summed expression for coexpression analyses. The data were filtered to remove cells with a mitochondrial ratio > 20%, less than 200 genes or 500 counts detected or that failed to meet a complexity score (ratio of genes to counts) cutoff of 80%. For each cell type, data had been collected from more than one foetus and thus the harmonise method in the Seurat package was used to correct for batch effects. Gene modules that contained *EP300* were identified and pairwise co-expression between *EP300* and all other TFs in a model was determined using both the Pearson and Spearman methods. As with bulk RNA-seq, *p* < 0.05 for either test was considered a true correlation.

### Cell Culture

H9 Human Embryonic Stem Cells (hESCs) (WiCell®, WA09), were maintained by diluting 1ml of cell suspension into 5 ml of Essential 8™(E8) media [Gibco™, A1517001] in vitronectin-coated (Gibco ™, A14700) 25ml flask (Thermo Scientific ™, 156367). The hESCs were differentiated into neurons using the dual SMAD inhibition protocol^28^, with modifications, for 28 days and the cells were grown as neurospheres from day 14-28. The cells were exposed to VPA throughout differentiation from day 0-28 and divided into 3 exposure groups of 300µM, 700µM and a control with no VPA exposure. Media and drugs were refreshed every two days.

### mRNA Isolation & RNA sequencing

mRNA was isolated using the RNeasy® Plus Mini Kit (QIAGEN®, 74134) and cDNA was synthesised using the materials provided in the Quantitect® Reverse Transcription Kit (QIAGEN®, 205311) and protocol. RNA was prepared using a NEBNext® Ultra II™ Library Prep Kit for Illumina® and was of good quality (Q30 >90%). Bulk RNA-sequencing on the Illumina® NovaSeq™ 6000 was undertaken by the Monash University Genomics Facility with PE150 at 30M reads/sample.

### *hESC* bioinformatics

The raw reads were aligned to human genome build GRCh38 and an in-house pipeline was applied for quality filtering and to construct a normalised count matrix. Gene expression was compared between unexposed cells and cells exposed to both VPA doses using the limma-zoom R package^29^ and genes with adjusted *p* < 0.01 were considered to be differentially expressed [Supplementary Figs. S10-11]. As with the public data both the Pearson and Spearman methods were applied to determine the correlation between *EP300* and all TFs on our list that were present in the data with *p* < 0.05 considered to be true correlation.

## Results

The study was approved by the Melbourne Health Human Research and Ethics Committee and written informed consent was obtained from all participants. The cohort comprised 250 mothers, 48 of whom had one or more affected pregnancies (cases). Sixty-six pregnancies were exposed to VPA as monotherapy or polytherapy, with 28 cases, and 184 were exposed to other ASMs, with 20 cases (Table 1, Supplementary Table S1). Following post-quality control filtering of the imputed data, 1,997,655 variants were analysed [Supplementary Table S2, Fig. S1].

**Table 1.** Affected pregnancies.

| **Mother** | **Birth defects** | **In Utero**<br>**ASMs**<br>**(mg/day)** |
| --- | --- | --- |
| AM21 | Cryptorchidism | VPA (800) |
| AM11 | Right renal hydronephrosis | VPA (600), LTG (100) |
| AM22 | Extra thumb | VPA (600) |
| AM2 | No bone in left thumb, tight fingers in both hands, and cardiac defects (VSD and ASD) | VPA (1000) |
| AM10 | Ventricular septal defects (bulbus cordis and cardiac septal closure) | VPA (1000), LTG (200) |
| AM12 | Dysmorphia | VPA (500) |
| AM13 | Two pregnancies: Retrognathia and hypertolism and hypospadias and induced TOP due to spina bifida and hydrocephalus | VPA (2000), LTG (150) |
| AM14 | Spina bifida and ureteric obstruction | VPA (1000) |
| AM15 | TOP due to anencephaly | VPA (UNK), CBZ (UNK), CLN (UNK) |
| AM16 | TOP due to anencephaly | VPA (400) |
| AM4 | Hypospadias | VPA (800), CLN (0.25) |
| AM17 | Bilateral deafness: left ear auditory nerve missing, right ear malformed cochlear | VPA (400) |
| AM3 | Club foot | VPA (1000), LEV (1500) |
| AM18 | TOP due to spina bifida | VPA (3000), TPM (75) |
| AM19 | TOP due to spina bifida | VPA (3000) |
| AM20 | Two pregnancies: patent foramen ovale, and cardiac defect and cleft lip and palate | VPA (500) |
| AM1 | Arnold-Chiari malformation, clinodactyly, cardiac defects (VSD and ASD significant, PDA small) | VPA (1500) |
| IM8 | Cleft palate | VPA (3000), TPM (500), PHT (350) |
| IM9 | Neural tube defect | VPA (1000) |
| IM10 | Club foot | VPA (500), LTG (25) |
| IM11 | Periauricular skin tag and brachicephalic skull | VPA (1500) |
| IM12 | Dysmorphic | VPA (1000) |
| IM13 | Limb defect (shortened arm) | VPA (1000), LTG (25), PHT (300), OXC (1800), CLB (10), TGB (30) |
| IM14 | Cleft palate | VPA (1500) |
| IM1 | Spinal deformity | VPA (1600) |
| IM2 | Arnold-Chiari, syringomyelia, hydrocephalus, developmental delay | VPA (1500), LTG (200) |
| IM3 | One child with hypospadias (two additional pregnancies with developmental delay/autism, VPA 2000 and 1200) | VPA (1500) |
| IM6 | Gastroschisis | VPA (800) |
| IM4 | Cardiac (heart murmur), umbilical hernia | LTG (500), CBZ (600) |
| IM5 | Skeletal (extra digits) | LTG (150), LEV (1500) |
| IM7 | Neural tube defect | CBZ (1200), LEV (3000) |
| AM23 | Oral (tongue tie) and hydronephrosis | LTG (200) |
| AM24 | Cryptorchidism | CBZ (600), LTG (100) |
| AM25 | Hypospadias | LEV (500) |
| AM26 | TOP due to defects (details unknown) | CBZ (800) |
| AM27 | Twin1 VSD and hypospadias, twin 2 VSD | CBZ (800) |
| AM28 | TOP due to bladder obstruction | LTG (200) LEV (500) |
| AM29 | Bilateral club feet | CBZ (1600) |
| AM30 | Hypospadias | LTG (100) |
| AM31 | VSD | LEV (1000) |
| AM32 | Tetralogy of Fallot | LEV (1500) |
| AM5 | Macrocephaly, dysmorphia and ankyloglossia | CBZ (1200) |
| AM8 | Cryptorchidism | PGB (450), TPM (300) |
| AM33 | Missing toe and syndactyly | LEV (1000) |
| Am34 | Bilateral club feet | LTG (200) |
| AM6 | Hypospadias | CBZ (500) |
| AM35 | Bicuspid aortic valve | LTG (200) |
| AM36 | ASD | OXC (1500), CLB (20) |

ASD\_ = Atrial Septal Defect, ASM= Antiseizure medication, CLB = Clobazam, CBZ = Carbamazepine, CLN = Clonazepam, LEV = Levetiracetam, LTG = Lamotrigine, OXC = Oxcarbazepine, PHT = Phenytoin, TGB = Tiagabine, TOP = termination of pregnancy, TPM = Topiramate, UNK = Unknown, VPA = valproic acid, VSD = Ventricular Septal Defect. All births were live births unless TOP is specified.

### Variant burden in genes associated with birth defects

The MalaCards database identified 1,525 autosomal genes associated with one or more of 32 birth defect types (Supplementary Table S3). The number of genes with a variant burden significantly different between cases and controls was higher in pregnancies exposed to VPA (either as monotherapy or polytherapy) compared to those exposed to other ASMs (Supplementary Table S4, OR 1·73 [95% CI 1.39 to 2.13], *p* = 2.25×10^-7^) and less than 2% (27/1525) of genes had a variant burden that was significantly different between cases and controls in both groups. The number of unique gene-birth defect type associations was higher for the VPA group than the other ASMs group (OR 1.56 [95% CI 1.32 to 1.84] *p* = 1.11×10^-7^) and all 32 birth defect types were implicated in the former but only 29 in the latter. For each of the six burden tests applied, none or less than 0.003% of random genes across 100 randomly sampled gene sets had a p value smaller than that obtained for birth defect-related genes [Supplementary Fig. S2].

### VPA-exposed Birth defect-hub (DB-hub)

When viewed as a protein-protein interaction network, 227 of encoded proteins formed a highly connected hub (BD-hub) representing genes associated with 31 different birth defect types (Fig. 1B, Supplementary methods, Supplementary Table S5). The difference in burden between cases and controls could be attributed to ‘common only’ variants for 36 genes and ‘rare only’ variants for 44 (Fig. 1A). The number of genes associated with specific birth defect types ranged from 1 to 88 (median =7), with microcephaly being the most frequently represented. The number of birth defect types each gene was associated with ranged from 1 to 10, with 9 genes contributing to 5 or more types (Fig. 2).

**Figure 1:**
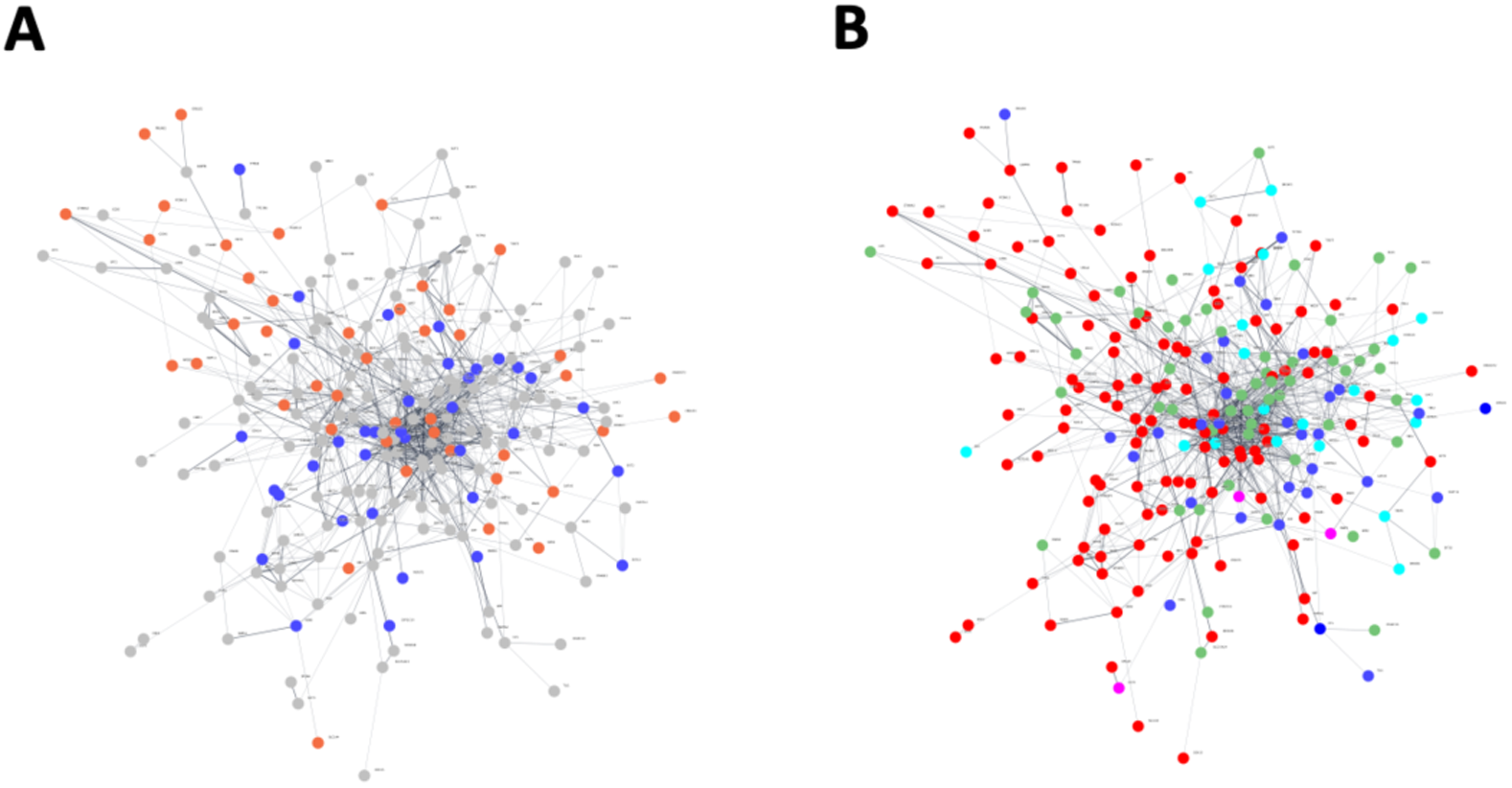
BD-hub comprising genes with significantly different variant burden between affected and unaffected VPA-exposed pregnancies. (**A**) **Variant burden:** for genes coloured in blue (n=36) the difference in burden between affected and unaffected pregnancies was attributed to ‘common only’ variants and those attributed to ‘rare only’ variants are shown in orange (44). (**B**) **Defect types**: Hub genes are coloured by specific birth defect types: red (*n* = 5) neural tube defects, anencephaly, encephalocele, microcephaly, and microphthalmia; dark blue (*n* =7) Atrial heart septal defect, atrioventricular septal defect, conotruncal heart malformations transposition of the great arteries dextro-Looped, heart septal defect, tetralogy of fallot and ventricular septal defect; pink (*n* = 3): biliary atresia, gastroschisis, and intestinal atresia; green (*n* = 10) diaphragmatic hernia, congenital, cleft palate isolated, craniosynostosis, cleft lip with or without cleft palate, clubfoot, fryns microphthalmia syndrome, microtia, omphalocele, and esophageal atresia; and light blue (*n* = 4) congenital anomalies of the kidney and urinary tract, renal hypodysplasia, cryptorchidism and hypospadias. Two conditions, atrioventricular septal defect and meningocele, are not shown as they were each assosiated with a single gene that was also implicated (and coloured) in other conditions.

**Figure 2.**
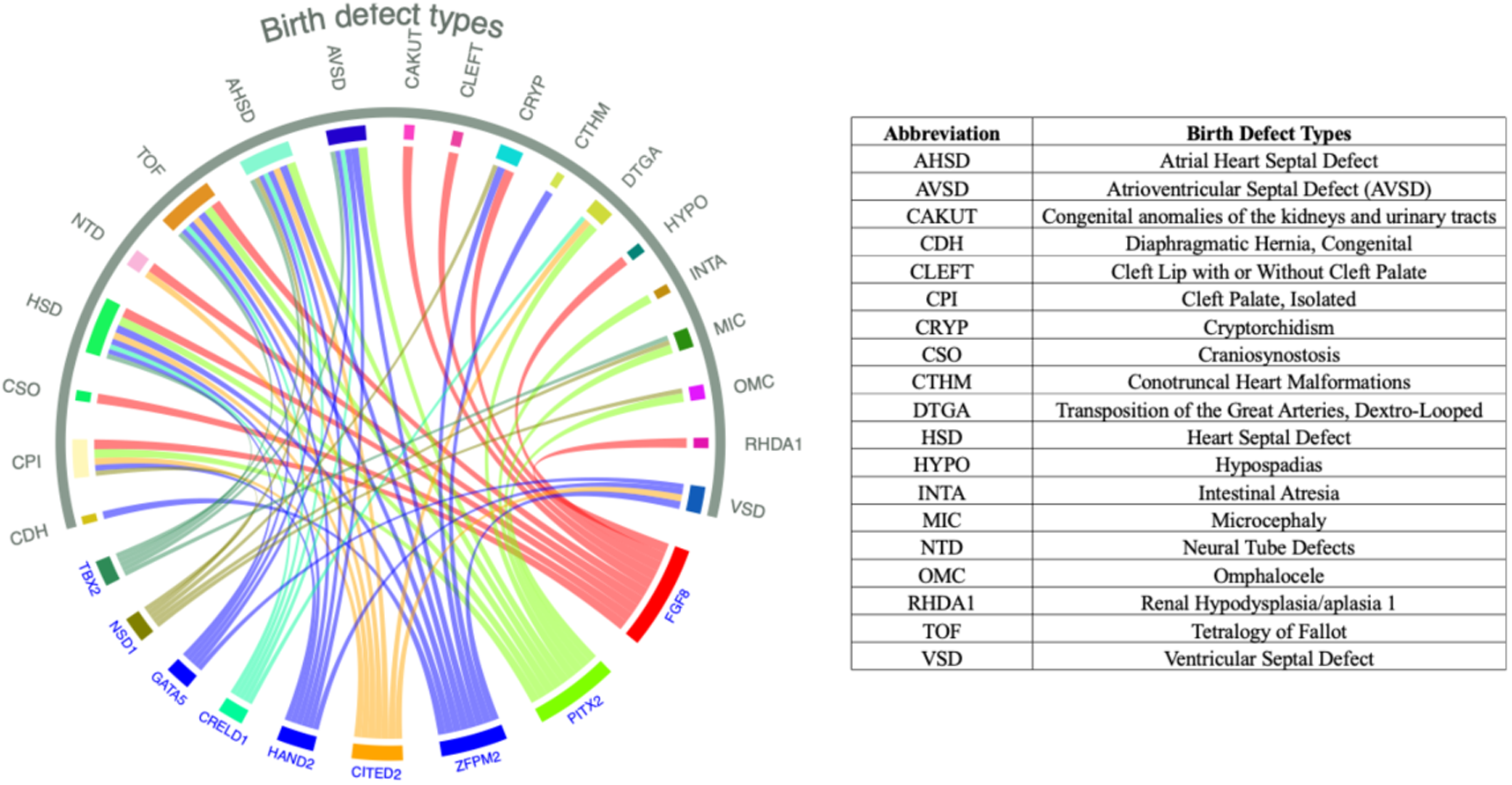
Association between genes within the VPA-exposed BD-hub and birth defect types. The ribbons represent the association between genes (blue labels bottom of circle) and specific birth defects abbreviations (green labels top of circle, full names in table) based on information from the Human Disease Database MalaCards.

Most variants within the VPA-exposed BD-hub genes lay within intronic (82%), downstream (6.92%) or upstream (4·59%) gene regions (Supplementary Table S6). This is in line with a comparison of SNP detection by two commercial exome capture platforms (Agilent and NimbleGen) which found that intronic variants were most represented among the roughly 25,000 to 40,000 SNPs identified by both platforms.^30^ Integration of eQTL data showed that most BD-hub genes (94%, 214/227) harbor variants that modify gene expression (*n* = 14,658, range per gene = 1 to 846, median = 32 [IQR 10.25 to 82]). A single variant can impact the expression of multiple genes including (but not always) the gene they fall within. The eQTL data indicated that the expression of 81% (183/227) of the BD-hub genes was dependent on variant(s) they or other genes within the hub harbour, inferring that the functional impact has relevance for birth defect biology. Sixty-two percent (9154/14,658) of the eQTL variants were predicted to impact the binding affinity of one or more TFs. The number of variants with both sources of functional evidence was lower in the BD-hub derived from non-VPA exposed pregnancies (*n* = 6,584) and across 100 randomly generated gene sets (maximum 2,219, median 1,096 [IQR 912 to 1,400], Supplementary Table S7).

### Integration of published differential expression data

The integration of differential expression data from multiple sources showed that 79% (180/227) of BD-hub genes had evidence of being dysregulated by VPA including 85% (45/53) of those with the highest degree (number of direct neighbours > 10) within the hub (Supplementary Fig. S4). The percentage of genes with evidence of VPA-induced dysregulation was lower 68% (90/132) in the BD-hub generated using data from non-VPA exposed samples, while the percentage of CBZ-induced dysregulated genes was slightly higher in the non-VPA exposed BD-hub (15%,20/132) than in the VPA-exposed BD-hub (12%, 28/227).

### Variant impact on TF-binding

Variants within genes in the BD-hub were predicted to modify the binding affinities of 359 TFs. The encoded proteins formed a densely connected network: the TF-hub. Highly connected genes within the TF-hub have master regulatory roles in stem cell pluripotency (*SOX2, POU5F1, NANOG* and *MYC*), and embryonic development (e.g. members of the GATA family (GATA1-4) that play a prominent role in regulating cardiac development and *PAX6* which is involved in the regulation of forebrain development).^31^ The most connected, linking to 42% (151/359) of all other TFs, was the acetyltransferase and transcriptional coactivator *EP300.* Five TFs had protein interaction evidence with 100 or more TFs in the network (*EP300, MYC, SOX2, JUN* and *FOS*). *FOS* and *JUN* are dimers of the AP-1 (activating protein-1) complex which is regulated through interactions with coactivators including *EP300*.^32^ The binding affinity of one or more of *EP300*, *FOS* and *JUN* was predicted to be modified by 465 variants that lie within 122 of the 227 (54%) genes in the BD-hub; linking disruption of this regulatory mechanism with 27 unique birth defect types [Supplementary Table S8].

The impact of genetic variation, measured by the number of variants predicted to impact on the binding affinity of any of the 50 most connected TFs, was higher for gene variants in the VPA-exposed BD-hub than the BD-hub derived from mothers exposed to other ASMs (Wilcoxon signed-rank test [95% CI 63.50 to 106.50], *p* = 1.1E-09), and variants within randomly generated sets of genes (all one sample t-test/ Wilcoxon signed-rank tests *p* <0.001, Supplementary Table S9, Fig. 3). In the TF-hub, 70% (249/359) of TFs were dysregulated by VPA exposure including 18/22 with a degree >70 (Supplementary Fig. S5), and 12.5% (45/359) had evidence of CBZ-dysregulation with 38 being dysregulated by both drugs.

**Figure 3.**
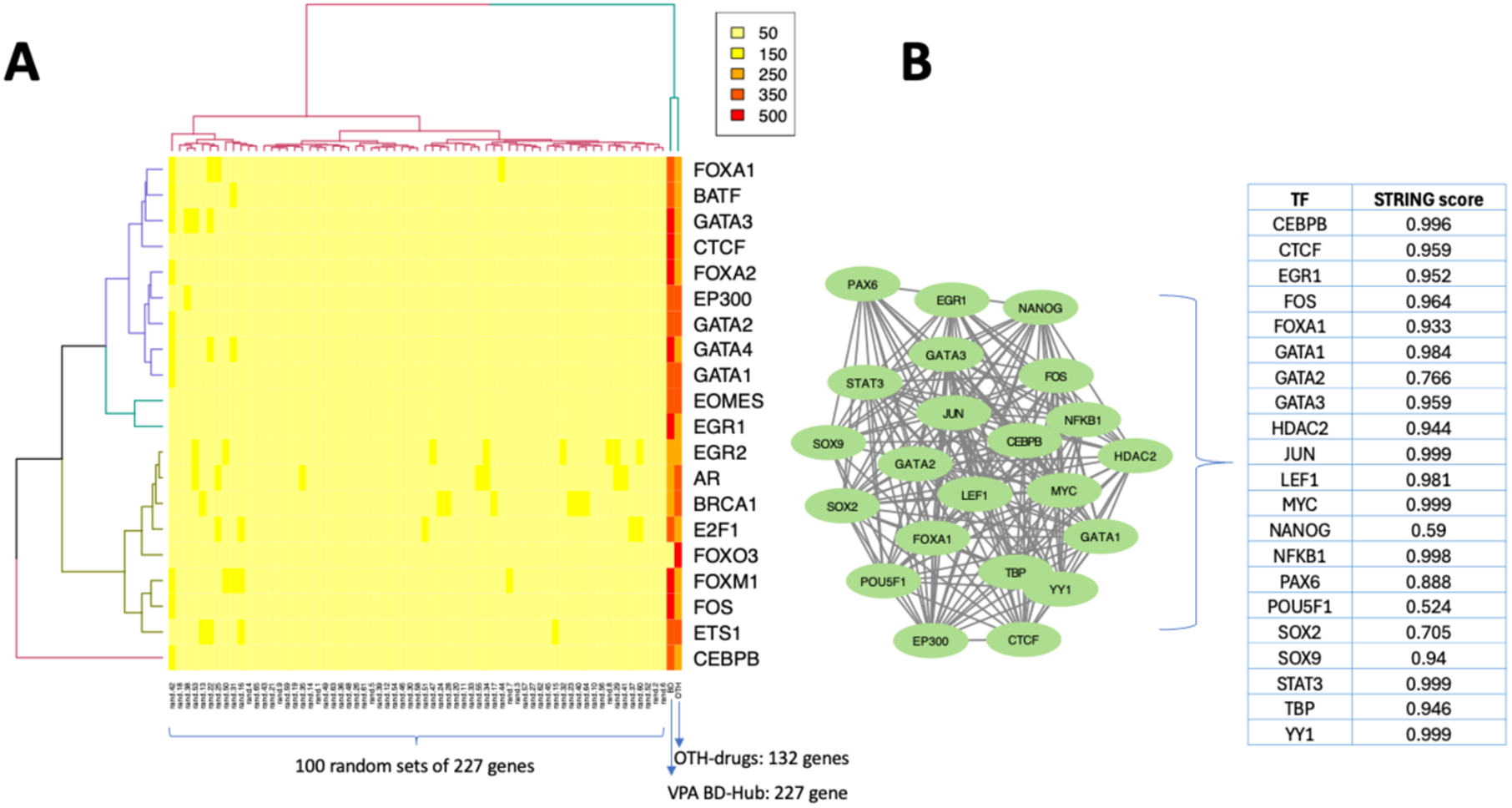
**(A) Variant burden.** Heatmap representing the predicted impact of variants on the binding affinity of the top 20 most highly connected TFs in the TF-hub. The rows represent TFs and the columns are gene sets. The first two columns on right-hand side of the x-axis represent the impact of variants within a gene hub derived from genes significantly different between cases and controls exposed to other ASMs, and variants within the VPA-exposed BD-hub. The remaining columns represent variants within 100 randomly generated gene sets. A stronger colour represents a higher number of variants predicted to modify a TF. **(B) *EP300* interactions.** Protein-protein interactions between *EP300* and other TFs within the TF-hub. Oval shapes represent genes encoding the proteins and the lines connecting genes are determined by STRING database scores (shown in table) which are ranked from 0 to 1 depending on how likely STRING judges an interaction to be true.

There was no statistical difference across the gene hubs in terms of overlap with a transcriptomic signature used to classify teratogenic compounds^27^ (53/227 of VPA-exposed, 34/132 of non-VPA exposed and 89/359 of the TF-hub genes, chi-squared test of homogeneity: x^2(2) = 0.29, *p* = 0.864), and little overlap in the genes implicated across the hubs. Of note, the TF-hub included Meis Homeobox 2 (*MEIS2*), one of the top four teratogenic signature genes (out of 2,757 analysed) that is differentially expressed by 10 different teratogenic compounds.

### High confidence variants

One hundred and thirty-five variants met our high-confidence filtering criteria. Comparison of the mean read depths (Wilcoxon signed-rank test, Supplementary Fig. S6) at the loci of high-confidence variants in VPA-exposed samples found no significant difference between Nextera and the Broad institute custom kit (*n* = 51 vs *n* =12 [95% CI −5.00 to 7.00], *p* = 0.85) or NimbleGen (*n* = 3, [95% CI −11.0 to 22.0], *p* = 0.865) or between NimbleGen and the Broad custom kit ([95% CI −9.99 to 13.99], *p* = 0.51). These variants were ranked based on the potential to disrupt one or more ‘master regulators’ (degree >70) identified within the TF-hub (Supplementary Table S10). The highest ranking ‘high confidence’ variant [chr11:49173538:T:C] lies within the intronic region of *FOLH1*, the gene encoding Folate Hydrolase 1, that is associated with neural tube defects^33^ and down-regulated by VPA in a cortical organoid model.^25^ The variant was detected by all exome capture kits (Supplementary Fig. S6). *FOLH1* met the significance cut-off for only one SKAT test, which evaluated the contribution of both common and rare variants (*p* = 0·02), but the frequency of this single variant was higher in VPA-exposed cases compared to controls (Empirical *p* = 0.007). Relevant to the homozygous reference genotype TT, the TC genotype has tissue and cell type-specific impact: reduced expression of *FOLH1* in arterial and nerve cells and tissues of the colon, esophagus and omentum; and increased expression of a brain specific transcript (RP11-707M1.1), across 6 different brain regions (Supplementary Fig. S7). The DeepBind scores predict that this variant modifies the binding of 15 TFs including 6 master regulators (*CEBPB, EP300, FOS, HDAC2, JUN* and *STAT3*), four of which have evidence of VPA-induced dysregulation (*EP300, FOS*, *JUN* and *STAT3*). Together, these observations suggested that both the genetic variant and prenatal VPA exposure could modify the activity of multi-protein complexes, with their combination potentially having an additive effect.

The overall spectra of high confidence variants suggested that genetic variation across multiple genes can converge to modify a specific TF protein complex. An example is the GATA family of TFs which comprises six proteins (GATA1-6) that play a critical role in the early stages of cell differentiation and organ development, involving tissues from all germ layers.^34^ The dysregulation of pleiotropic activity and tissue specific regulation of this type of multi-protein complex might explain the wide range of VPA-associated birth defects (Fig. 4, Supplementary Table S8).

**Figure 4:**
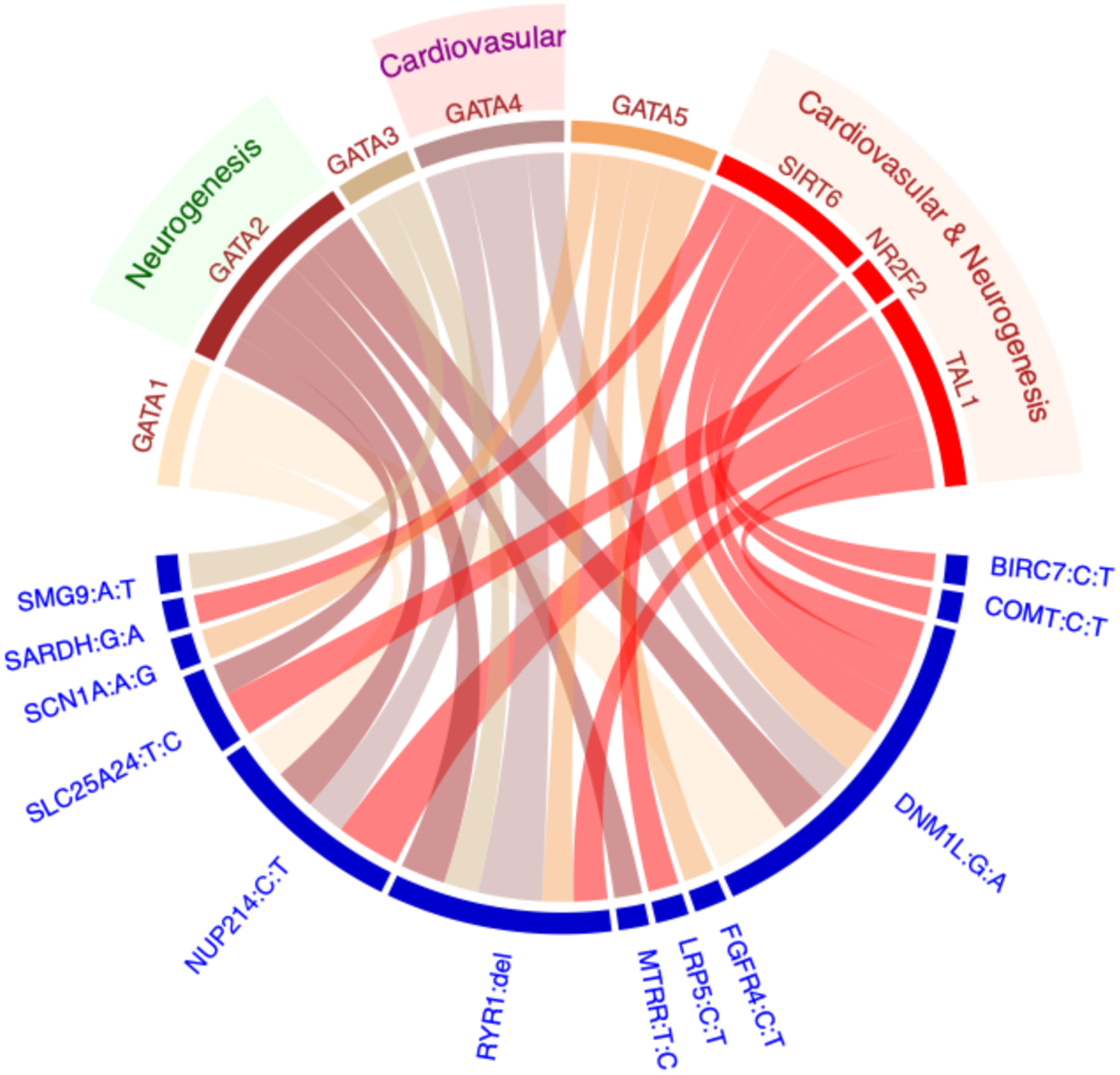
Transcription factors as a link between genetic variation and birth defects. The ribbons in the circos plot link 12 gene:variants (blue text) to TFs with tissue-specific effects (*GATA1-5*, *NR2F2*, *SIRT6* and *TAL1*). The thickness of a connection is proportional to the score generated by DeepBind which represents the difference in predicted binding affinity between wildtype and variant-containing alleles.

### *EP300*-TF coexpression reference map

*EP300* was significantly coexpressed with 91% (2, 2144/2,367) of TFs tested in bulk RNA-seq data from the BrainSpan repository representing cortical and subcortical structures across the full course of human brain development [Supplementary Fig. S8A]. In scRNA-seq data, 238 coexpressed gene modules involving TFs were detected across two tissue and 10 cell types (Supplementary Table S11, Fig. S8B). Of these, 12% (29/238) contained *EP300*. Within these modules *EP300* was coexpressed with 86% (2,011/2,347) of TFs tested, and 83% (1812/2176) of coexpressed pairs were found in both bulk and single cell settings. Overall, the number of times a unique coexpression relationship was observed ranged from 1 to 18 out of a possible 25 biological settings, median = 9, with 1,053 observed in 10 or more unique settings. For 71% (1662/2341) of TFs profiled, the direction of correlation differed across biological settings with this being observed more across tissues (55%,1206/2176) as compared to cell types (26%, 535/2011).

### Impact of VPA exposure on *EP300* coexpression

Gene expression data from unexposed hESCs, showed *EP300* to be coexpressed with 422 TFs and 99% (417/422) of these relationships were observed in one or more settings in the reference map (Fig. 5A, range 1 to 18, median 10). Similarly, almost all coexpressed pairs observed in cells exposed to VPA were present in the reference (99% (223/226) at 300µM and 99% (452/456) at 700µM, Fig 5A). In contrast, in our hESC model only 32% (134/422) of the coexpressed pairs observed in unexposed cells were present in cells exposed to VPA (38 at 300µM, 109 at 700µM, with 13 observed at both doses, Fig. 5B). Among the 288 TFs for which coexpression with *EP300* was lost, just over half (58%, 166/288) were also differential expressed at one or both VPA does. Similarly, only 46% (224/489) of pairs observed in VPA-exposed cells only were also dysregulated at one or both doses. There was no significant difference in the level of *EP300* mRNA following exposure (adjusted *p* = 0.334 at 300µM, adjusted *p* = 0.706 at 700µM). These finding indicate that the impact of VPA on the activity of regulatory mechanisms is likely not fully exposed by studies that focus on differential expression alone.

**Figure 5:**
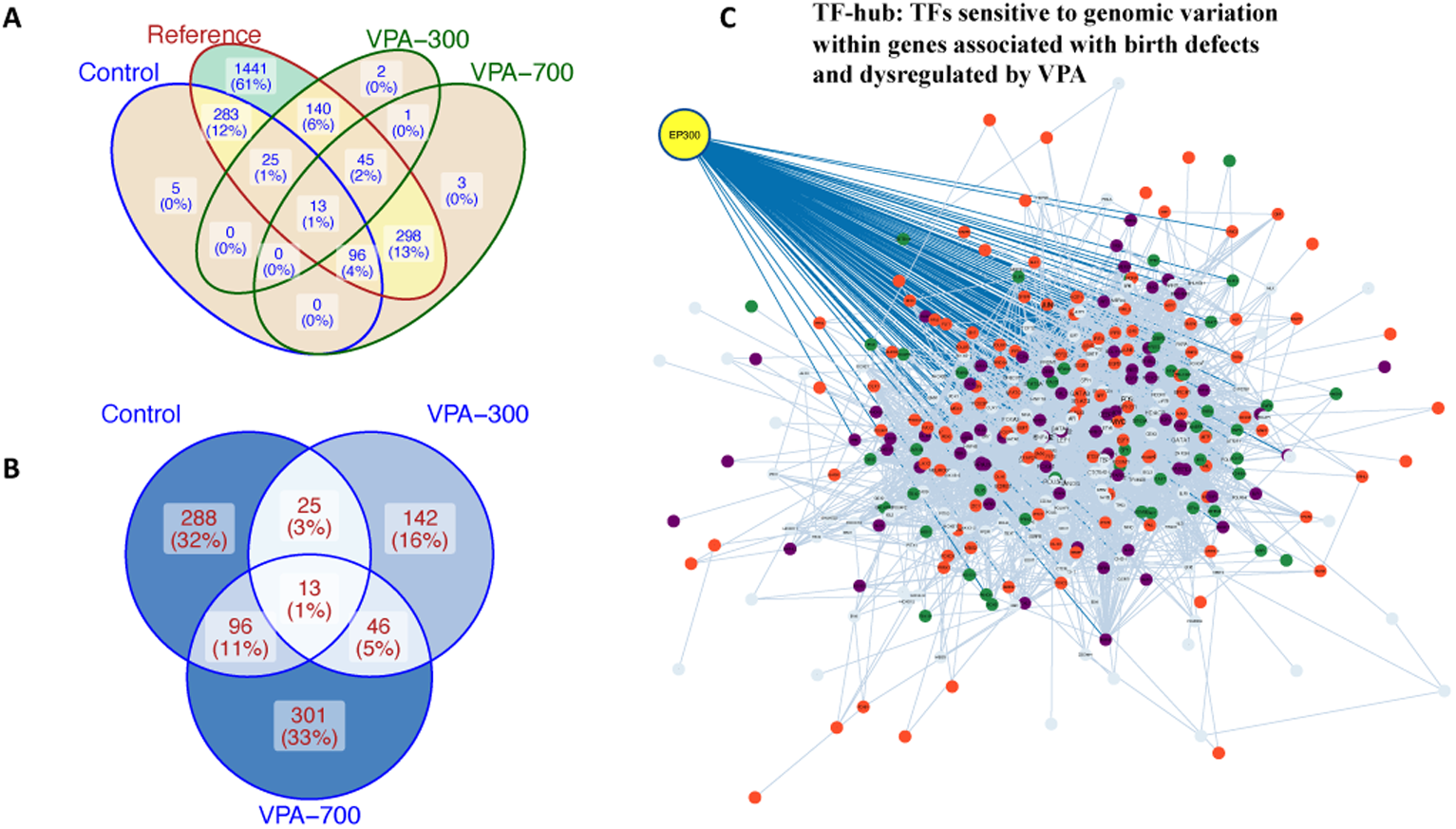
Results from coexpression analyses: **(A) Comparison between reference and hESC model**: venn diagram showing good overlap in correlated expression of *EP300* with other TFs between those observed in public reference data and data obtained from our hESC exposure model. **(B) Comparison between exposure groups:** venn diagram showing the extent of differences in *EP300* correlated expression between unexposed and VPA exposed cells. (**C**) **Differential expressed and coexpressed TFs in the TF-hub:** overlay of results comparing unexposed and VPA-exposed at either dose found 64% (229/359) of the TFs in the hub were either both differentially expressed and differentially coexpressed with *EP300* (*n* = 66, purple nodes), differentially coexpressed with *EP300* only (*n* = 48, green nodes) or differentially expressed only *n* =114, orange nodes).

## Discussion

The molecular mechanisms underlying VPA anatomical and behavioural teratogenicity remain poorly understood. A genetic predisposition among mothers could explain why not all pregnancies are affected, however, to date genetic biomarkers have been elusive. In this work, we considered whether genetic variation that disrupts gene regulatory activity of TFs might contribute to variation in both teratogenic effects and risk to offspring of mothers taking VPA during pregnancy. As would be expected, given that TFs rarely act in isolation, we found that variants within different genes potentially have converging effects on a single regulatory mechanism, and that a single variant could disrupt the binding of multiple TF complexes.

The potential sensitivity of *EP300*-mediated TF complexes to genetic variation was an important, novel finding in this study. Our expression correlation analyses in hESCs indicate that the activity of *EP300*-related TF complexes, differs considerably between unexposed cells and VPA exposed cells, while correlated pairs in all exposure settings have high overlap with those in the reference map. The strong overlap in exposed cells and the reference suggest that VPA alters regulatory programs specific to neuronal differentiation, by activating alternative valid, but context inappropriate, signaling pathways. Disruption of this regulatory mechanism could underlie the high levels of differential expression reported in previous studies but remained elusive as not all the TFs found to be differentially coexpressed were simultaneously differentially expressed.

The *EP300* gene encodes p300, a member of the cAMP response element-binding protein (CREB)- binding protein family (CBP). p300 is critical to embryogenesis being embryonic lethal in nullizygous and heterozygous mice.^35^ The CBP family of proteins have histone acetyltransferase (HAT) activity. We found no significant difference in the level of *EP300* mRNA following exposure to VPA in our cell model. *In utero* exposure in a mouse model of neural tube defects found p300 mRNA, but not protein levels, to be downregulated at 1 and 3 hours post-exposure to VPA in GD9 mouse embryos^36^ while both mRNA and protein levels of p300 were decreased following exposure in a mouse cell line.^37^ Among embryos exposed or not to VPA, the level of p300 mRNA and protein expression in those with closed or open neural tubes were not significantly different.^36^ Changes in p300 activation may be more important for pathogenicity than cellular protein levels. The p300 enzyme is activated by short-chain fatty acids (SCFAs) acetate, propionate, and butyrate, that are produced by the gut microbiome.^38^ Whether the presence of VPA, also a SCFA, can aberrantly activate or otherwise modify the function of p300 is worthy of further investigation.

AP-1 subunits *FOS* and *JUN* were among the most connected TFs in the TF-hub. The genomic occupancy of *EP300* is enriched at AP-1 hotspots,^32^ and we found that over half the genes in the BD-hub, associated with 27 different birth defect types, harboured variants predicted to impact on the binding of *FOS, JUN* or *EP300*. The AP-1 complex has been identified as a necessary mediator of both VPA-perturbation of neural crest differentiation and rescue by co-culture with rapamycin, in a spinal cord organoid model of teratogenicity.^39^ Whether the collective burden across these variants, or birth defect type-specific variants among them, determine risk of teratogenicity more broadly is worthy of investigation in larger patient cohorts.

We identified a candidate variant within *FOLH1* that was predicted to impact the binding activity of 15 TFs, including p300. While the functional consequence of intronic variants may be entirely distinct from the function of the protein encoded by the gene they fall within, the eQTL data show that this variant impacts the expression of the *FOLH1* gene, reducing expression in tissues of the colon and esophagus. Perturbation of TF binding by this variant may be a plausible explanation for the difference in expression. *FOLH1* encodes a transmembrane glycoprotein that acts as a glutamate carboxypeptidase on substrates including the nutrient folate. In a rat model of hypertension, Folh1 was expressed in the small intestine and animals with mutated Folh1 had reduced folate clearance and lowered plasma cysteine and homocysteine levels.^40^ It is interesting to speculate that individuals who carry this variant may have a predisposition to folate deficiency, due to impaired folate processing, that is exacerbated by VPA treatment, or reduced capacity to benefit from folate supplementation. It is noteworthy that folate supplementation has not been found to protect against VPA-associated neural tube defects or other teratogenic birth defects in the same way it has for the general population.^41^

The use of whole-exome, rather than whole-genome, sequence data is a limitation of this study. However, it has been shown that enhancers, with tissue-specific activity are enriched in intronic gene regions,^20^ while genes that are ubiquitously expressed, such as housekeeping genes, are controlled by intergenic enhancers.^20^ The relatively small sample size in this study is also a limitation and larger cohorts with VPA-exposed pregnancies are needed to tease out specific regulatory mechanisms that are sensitive to VPA and to evaluate neurodevelopmental risk in addition to structural birth defects. We acknowledge that the large number of TF binding sites implicated likely include spurious calls due to limitations of the DeepBind tool used to detect them. By coupling these predictions to more robust eQTL data, we helped to overcome this limitation. While we cannot yet conclude with certainty that VPA modifies the binding of TFs, or their *EP300*-dependent regulation during embryogenesis, our findings align with the broader literature and provide a proof-of-concept that this type of genetic variation could contribute to VPA teratogenicity and warrants further investigation.

In conclusion, by contextualizing maternal exome variants to structural birth defect biology, and evaluating impact on TF-driven gene regulation, we find evidence in support of a putative mechanism for VPA anatomical teratogenicity that involves the “master gene transcription regulator”, p300. Our findings support the notion that variant-impacted TF activity could explain variation in both risk, and VPA-associated birth defect types, in women treated during pregnancy.

### Data Sharing Statement

In accordance with the ethics requirements of the Raoul Wallenberg Australian Pregnancy Register of Antiepileptic Drugs [Reference Number: HREC/11/MH/282], the exome data used in this study cannot be shared in a public repository. RNA-seq data from our hESC assay are available from Gene Expression Omnibus (GEO) Accession Number GSE290300.

### Contributors

TJOB, AA, PP, IES, SFB, PK, JC and FJEV conceived the study. AA designed and performed the bioinformatics analyses with assistance from EV. EV, DS, and AAB generated hESC models. TOB, JC, FJEV, DJ, AH, JG and MT were involved in data collection and project administration. SMS, TT, DB, EP, MMR, AR, LL, MPC, EZ, EC, AR, JC recruited the patients and/or were involved in establishing the pregnancy pharmacogenomic collaboration. TJOB, SMS and JC lead the funding applications. All authors contributed to writing, review and editing the manuscript and all approved the final version.

## Data Availability

RNA-seq data from our hESC assay are available from Gene Expression Omnibus (GEO) Accession Number GSE290300

https://www.ncbi.nlm.nih.gov/geo/query/acc.cgi?acc=GSE290300

## Acknowledgements

We thank all individuals who took part in the study, and Dr Zhibin Chen for his advice on statistical analysis.

## References

1. Marson A, Burnside G, Appleton R, Smith D, Leach JP, Sills G, et al. The SANAD II study of the effectiveness and cost-effectiveness of valproate versus levetiracetam for newly diagnosed generalised and unclassifiable epilepsy: an open-label, non-inferiority, multicentre, phase 4, randomised controlled trial. The Lancet. 2021;397(10282):1375–86.

2. Marson AG, Al-Kharusi AM, Alwaidh M, Appleton R, Baker GA, Chadwick DW, et al. The SANAD study of effectiveness of valproate, lamotrigine, or topiramate for generalised and unclassifiable epilepsy: an unblinded randomised controlled trial. The Lancet. 2007;369(9566):1016–26.

3. Gotlib D, Ramaswamy R, Kurlander JE, DeRiggi A, Riba M. Valproic acid in women and girls of childbearing age. Current psychiatry reports. 2017;19:1–7.

4. Natale G, Fini E, Calabrò PF, Carli M, Scarselli M, Bocci G. Valproate and lithium: Old drugs for new pharmacological approaches in brain tumors? Cancer Lett. 2023;560:216125.

5. GOV.UK. Valproate (Belvo, Convulex, Depakote, Dyzantil, Epilim, Epilim Chrono or Chronosphere, Episenta, Epival, and Syonell): new safety and educational materials to support regulatory measures in men and women under 55 years of age. In: Agency MaHpR, editor. UK2024.

6. Tomson T, Battino D, Bonizzoni E, Craig J, Lindhout D, Perucca E, et al. Dose-dependent teratogenicity of valproate in mono- and polytherapy An observational study. Neurology. 2015;85(10):866–72.

7. Vajda FJE, Hitchcock A, Graham J, Solinas C, O’Brien TJ, Lander C, et al. Foetal malformations and seizure control: 52 months data of the Australian Pregnancy Registry. Eur J Neurol. 2006;13(6):645–54.

8. Cohen JM, Alvestad S, Cesta CE, Bjørk MH, Leinonen MK, Nørgaard M, et al. Comparative safety of antiseizure medication monotherapy for major malformations. Ann Neurol. 2023;93(3):551–62.

9. Christensen J, Grønborg TK, Sørensen MJ, Schendel D, Parner ET, Pedersen LH, et al. Prenatal valproate exposure and risk of autism spectrum disorders and childhood autism. JAMA. 2013;309(16):1696–703.

10. Hernández-Díaz S, Straub L, Bateman BT, Zhu Y, Mogun H, Wisner KL, et al. Risk of autism after prenatal topiramate, valproate, or lamotrigine exposure. N Engl J Med. 2024;390(12):1069–79.

11. Meador KJ, Baker GA, Browning N, Clayton-Smith J, Combs-Cantrell DT, Cohen M, et al. Cognitive function at 3 years of age after fetal exposure to antiepileptic drugs. N Engl J Med. 2009;360(16):1597–605.

12. Meador KJ, Baker GA, Browning N, Cohen MJ, Bromley RL, Clayton-Smith J, et al. Fetal antiepileptic drug exposure and cognitive outcomes at age 6 years (NEAD study): a prospective observational study. The Lancet Neurology. 2013;12(3):244–52.

13. Vajda FJE, O’Brien TJ, Lander CM, Graham J, Roten A, Eadie MJ. Teratogenesis in repeated pregnancies in antiepileptic drug-treated women. Epilepsia. 2013;54(1):181–6.

14. Tomson T, Battino D, Bonizzoni E, Craig J, Lindhout D, Sabers A, et al. Dose-dependent risk of malformations with antiepileptic drugs: an analysis of data from the EURAP epilepsy and pregnancy registry. Lancet Neurol. 2011;10(7):609–17.

15. Lundberg YW, Cabrera RM, Greer KA, Zhao J, Garg R, Finnell RH. Mapping a chromosomal locus for valproic acid-inducedexencephaly in mice. Mamm Genome. 2004;15:361–9.

16. Vajda F, O’Brien T, Graham J, Hitchcock A, Perucca P, Lander C, et al. Specific fetal malformations following intrauterine exposure to antiseizure medication. Epilepsy Behav. 2023;142:109219.

17. Andrey G, Mundlos S. The three-dimensional genome: regulating gene expression during pluripotency and development. Development. 2017;144(20):3646–58.

18. Tseng C-C, Wong M-C, Liao W-T, Chen C-J, Lee S-C, Yen J-H, et al. Genetic variants in transcription factor binding sites in humans: triggered by natural selection and triggers of diseases. Int J Mol Sci. 2021;22(8):4187.

19. Nishizaki SS, Ng N, Dong S, Porter RS, Morterud C, Williams C, et al. Predicting the effects of SNPs on transcription factor binding affinity. Bioinformatics. 2020;36(2):364–72.

20. Borsari B, Villegas-Mirón P, Pérez-Lluch S, Turpin I, Laayouni H, Segarra-Casas A, et al. Enhancers with tissue-specific activity are enriched in intronic regions. Genome Res. 2021;31(8):1325–36.

21. Guo Y, Long J, He J, Li C-I, Cai Q, Shu X-O, et al. Exome sequencing generates high quality data in non-target regions. BMC Genomics. 2012;13(1):1–10.

22. Vajda F, Lander C, O’brien T, Hitchcock A, Graham J, Solinas C, et al. Australian pregnancy registry of women taking antiepileptic drugs. Epilepsia. 2004;45(11):1466-.

23. Perucca P, Anderson A, Jazayeri D, Hitchcock A, Graham J, Todaro M, et al. Antiepileptic Drug Teratogenicity and De Novo Genetic Variation Load. Ann Neurol. 2020;87(6):897–906.

24. Feleke R, Jazayeri D, Abouzeid M, Powell KL, Srivastava PK, O’Brien TJ, et al. Integrative genomics reveals pathogenic mediator of valproate-induced neurodevelopmental disability. Brain. 2022;145(11):3832–42.

25. Cui K, Wang Y, Zhu Y, Tao T, Yin F, Guo Y, et al. Neurodevelopmental impairment induced by prenatal valproic acid exposure shown with the human cortical organoid-on-a-chip model. Microsystems & nanoengineering. 2020;6(1):1–14.

26. Schulpen SHW, Pennings JLA, Piersma AH. Gene Expression Regulation and Pathway Analysis After Valproic Acid and Carbamazepine Exposure in a Human Embryonic Stem Cell-Based Neurodevelopmental Toxicity Assay. Toxicol Sci. 2015;146(2):311–20.

27. Cherianidou A, Seidel F, Kappenberg F, Dreser N, Blum J, Waldmann T, et al. Classification of developmental toxicants in a human iPSC transcriptomics-based test. Chem Res Toxicol. 2022;35(5):760–73.

28. Chambers SM, Fasano CA, Papapetrou EP, Tomishima M, Sadelain M, Studer L. Highly efficient neural conversion of human ES and iPS cells by dual inhibition of SMAD signaling. Nat Biotechnol. 2009;27(3):275–80.

29. Ritchie ME, Phipson B, Wu D, Hu Y, Law CW, Shi W, et al. limma powers differential expression analyses for RNA-sequencing and microarray studies. Nucleic Acids Res. 2015;43(7):e47-e.

30. Asan n, Xu Y, Jiang H, Tyler-Smith C, Xue Y, Jiang T, et al. Comprehensive comparison of three commercial human whole-exome capture platforms. Genome Biol. 2011;12:1–12.

31. Georgala PA, Carr CB, Price DJ. The role of Pax6 in forebrain development. Dev Neurobiol. 2011;71(8):690–709.

32. Seo J, Koçak DD, Bartelt LC, Williams CA, Barrera A, Gersbach CA, et al. AP-1 subunits converge promiscuously at enhancers to potentiate transcription. Genome Res. 2021;31(4):538–50.

33. Molloy AM, Pangilinan F, Brody LC. Genetic risk factors for folate-responsive neural tube defects. Annu Rev Nutr. 2017;37:269–91.

34. Tremblay M, Sanchez-Ferras O, Bouchard M. GATA transcription factors in development and disease. Development. 2018;145(20):dev164384.

35. Yao T-P, Oh SP, Fuchs M, Zhou N-D, Ch’ng L-E, Newsome D, et al. Gene dosage– dependent embryonic development and proliferation defects in mice lacking the transcriptional integrator p300. Cell. 1998;93(3):361–72.

36. Shafique S, Winn LM. Role of Cbp, p300 and Akt in valproic acid induced neural tube defects in CD-1 mouse embryos. Reprod Toxicol. 2020;95:86–94.

37. Lamparter CL, Winn LM. Valproic acid exposure decreases Cbp/p300 protein expression and histone acetyltransferase activity in P19 cells. Toxicol Appl Pharmacol. 2016;306:69–78.

38. Thomas SP, Denu JM. Short-chain fatty acids activate acetyltransferase p300. Elife. 2021;10:e72171.

39. Pietrogrande G, Shaker MR, Stednitz SJ, Soheilmoghaddam F, Aguado J, Morrison SD, et al. Valproic acid-induced teratogenicity is driven by senescence and prevented by Rapamycin in human spinal cord and animal models. Mol Psychiatry. 2024:1–13.

40. Šilhavý J, Marková I, Hüttl M, Malínská H, Kazdová L, Liška F, et al. Dissecting the role of Folr1 and Folh1 genes in the pathogenesis of metabolic syndrome in spontaneously hypertensive rats. Physiol Res. 2018;67(4):657–62.

41. Vajda FJ, O’Brien TJ, Graham JE, Hitchcock AA, Perucca P, Lander CM, et al. Folic acid dose, valproate, and fetal malformations. Epilepsy Behav. 2021;114:107569.

