## Supplementary methods for "Transcription factor binding and individual genetic risk for valproate teratogenicity"

### Data pre-processing

Data were available for 280 women with epilepsy (179 recruited in Australian and 101 through EpiPGX). Exome sequencing had been carried out through either the Broad Institute (Illumina HiSeq 2000 with Illumina TruSeq Exome Enrichment kit, or Broad custom kit), the Institute for Genomic Medcine, Columbia University in New York (Illumina HiSeq 2500 with Exome Kit version 3.0 (Roche NimbleGen) or deCODE genetics (Illumina HiSeq 2500 with Illumina Nextera Rapid Capture Expanded Exome kit).

The raw reads had been aligned either to the previous (GRCh37, hg19) or current (GRCh38, hg38) human genome build. The former were converted back to fastq files and realigned to the current genome build. Variant calling was undertaken using the Genome Analysis Toolkit (GATK, v.4.1.2) HaplotypeCaller in GVCF mode, yielding individual gVCF files that were subsequently used for joint-genotyping to generate a cohort variant call set. Variant quality scores were adjusted using the variant quality score recalibration method, with truth tranche scores between 99.9 and 100 considered false positives.

Two individuals were removed due to missing drug information. Pre imputation QC of samples included removing those with missingness > 5% ($n$ =14), with ambiguous or non-matching genetically imputed sex ($n$ = 3), heterozygous/homozygous ratio > 3 standard deviations from the mean ($n$ = 2), one of each pair found to be cryptically related (n=3) and one population outlier. Variants with missingness > 2% or that violated Hardy-Weinberg Equilibrium (HWE, p<10-6) were removed ($n$ = 47,277,056) and only autosomal variants with a minor allele frequency greater than 0.01 were considered for imputation (n=239,461). Imputation was undertaken using the Michigan imputation server with whole-genome sequencing-based apps@1000g-phase3-low@1.0.0 reference panel (5,008 haplotypes from 26 populations). Post imputation, dosage files were filtered to remove variants with estimated imputation accuracy (R-square) < 0.9 or violation of HWE or MAF < 0.01. Three heterozygous/homozygous ratio outliers were also removed. Post QC 250 individuals (48 cases, 202 controls) were available for analyses.

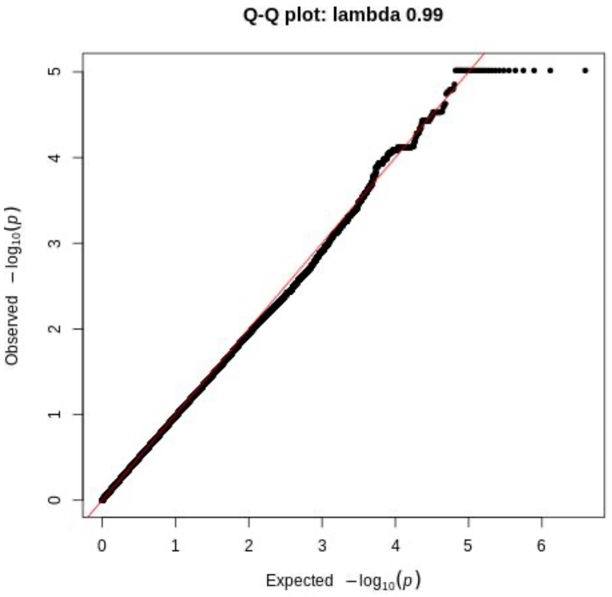


**Supplementary Figure S1:** QQ plot for post-imputation data derived from 1,997,655 variants and 150 individuals (48 cases, 202 controls).

**Gene set analysis**

The MalCards[1] resource was used to extract genes associated with 'structural birth defects' and 'genitourinary birth defects' and yielded 2,404 genes. The genomic positions for the genes were obtained using biomart in R and only autosomal genes (1,591) were retained for subsequent analyses. A total of 1,525 genes harboured at least one variant in at least one mother in the study cohort. Genes within this set were associated with one or more of 32 different birth defect types [range = 2-16, median =1, Supplementary Tables S2-S3]. The plink software (version 1.9) was used to generate gene level variant sets.

Gene set analysis was undertaken for women exposed to all drugs and subsets of women exposed to VPA in mono- or polytherapy and other drugs. Six burden tests available within the SKAT R library were applied to each gene set: SKAT.SSD.All, SKATBinary_Robust.SSD.All with method=SKAT and method=SKATO, SKAT_CommonRare.SSD.All with test.type="Common.Only", Rare.Only" or unspecified (both). A cut-off of $p < 0{\cdot}05$ for any test, was used to determine gene-birth defect type associations, within each study cohort.

**Construction of birth defect protein-protein interaction (PPI) networks (BD-hubs)**

The STRING database App within the Cytoscape software (version v3.10.2) was used with default settings (Confidence cut-off 0.4, max additional interactions=0, use smart delims ticked, load enrichment data unticked) to generate protein-protein interaction (PPI) networks for proteins encoded by significant birth defect-related genes (VPA-exposed cases vs controls, non-VPA cases vs. controls). The encoded proteins were represented as nodes and the lines joining nodes denoted interactions (based on information contained in the STRING database). The resulting networks were trimmed to include genes/proteins that linked to at least one other in the network [Supplementary Table S3, Fig. 1A-B].

**Random gene sets**

To establish the specificity of signal at the gene set level we used randomly generated gene sets. For each SKAT test, we compared P values for gene sets associated with birth defects against those generated from 1000 randomly selected sets, each comprising 44 genes (the median number of genes in gene sets based on genes associated with specific birth defects).

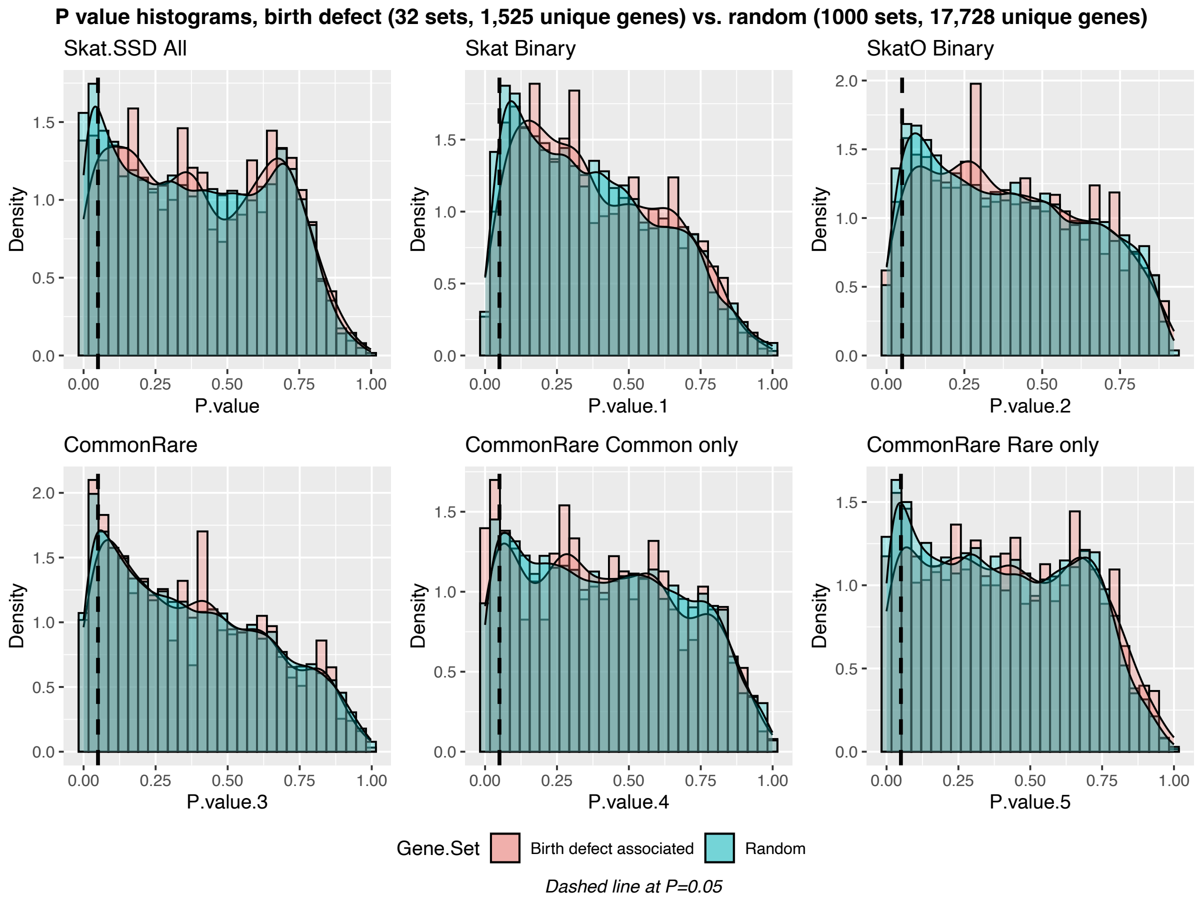


**Supplementary Figure S2: Distribution of SKAT test P values.** Histogram plots of gene level P values obtained using different SKAT tests for genes in the Birth defect sets (red) and random sets (green). For the Skat CommonRare test, none of the genes in the random sets had a P value smaller than the smallest observed across genes associated with birth defects. For the remaining SKAT tests, between 10 to 74 random genes had *p* values below those observed in birth defect-related genes [Skat SSD = 10/25445 ($p$ = 0.0003), Skat Binary = 74/25445 ($p$ = 0.0029), SkatO = 23/25445 ($p$ = 0.0009), CommonRare Common only = 24/25445 ($p$ = 0.0009) and CommonRare Rare only =13/25445 ($p$ = 0.0005).

**DeepBind tool**

The DeepBind algorithms had been trained using data from *in vitro* high-throughput SELEX (HT-SELEX) experiments for TF–DNA binding and the Chromatin immunoprecipitation (ChIP) followed by high-throughput DNA sequencing (ChIP-seq) approach for mapping the genomic location of transcription-factor binding and histone modifications in living cells, publicly available through the ENCODE project[2]. The functional consequence of variants was determined by comparing predicted binding affinity scores generated by DeepBind for the reference and variant-impacted DNA sequences at TF binding sites. A difference in scores close to zero indicated no functional consequence while scores ≤ -2 or > 2 were considered evidence of decreased (-ve) or increased (+ve) TF binding affinity. This cutoff was based on an empirical cumulative distribution function plot of the distribution of scores generated for all variants identified across all individuals in the cohort [Supplementary Fig. S3].

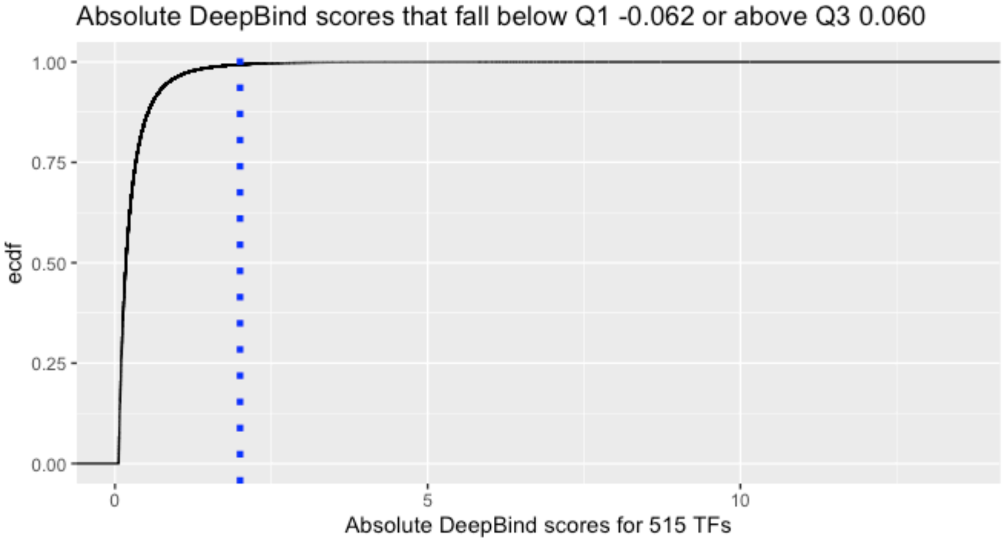


**Supplementary Figure S3: DeepBind scores***:* An empirical cumulative distribution function plot of DeepBind scores that fall outside the interquartile range. The scores represent variants that were observed in one or more individuals in the study. The majority centre around 0 indicating that they would have no impact on transcription factor binding (range -13.03 – 13.48, Median 0, Mean -0.002). A cutoff of absolute 2 was used to select those likely to disrupt binding.

**Construction of TF protein-protein interaction (PPI) network (TF-hub)**
368 TFs impacted by one or more variants within genes in the BD-hub that were also implicated in eQTL data were used to construct a PPI network using the Cytoscape software and information from the STRING database. No match was found for*, HINFP1* and *SIN3AK20* and of the remining 366 genes, 7 did not connect to any others and were excluded, leaving a highly connected hub with 359 nodes and 4,253 edges.

**Integration of evidence from transcriptomic studies**
In vivo[3] and in vitro models[4, 5] of VPA-induced teratogenicity show that high numbers of genes are differentially expressed under exposure to the drug[6]. Integration of VPA-dysregulated genes showed the log fold change (logFC) and number of BD-hub genes dysregulated was higher in data from a human forebrain organoid model of autism risk (45%, logFC range = 2.72 to 4.54) as compared to a rat model of VPA-induced neurodevelopmental disability[3] (18%, logFC range = -0.87 to 0.51), Supplementary Fig. S4C:D. Overlay of data from a human embryonic stem cell-test[6] of neurodevelopmental toxicity identified BD-hub genes that are dysregulated at one, seven or both days post VPA exposure (Supplementary Fig. S4B).

**A**

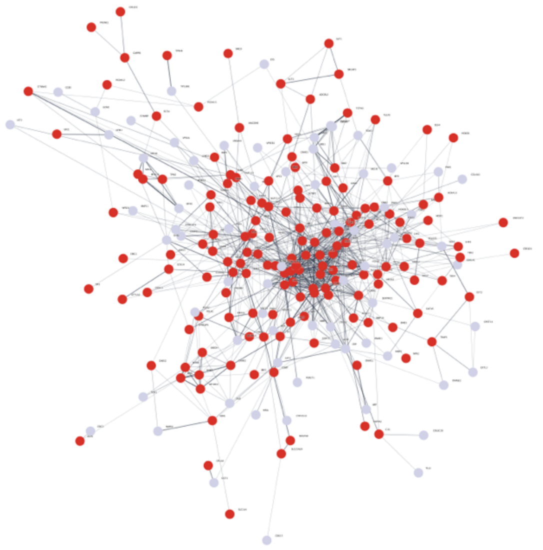

**B**

| Gene | Degree | Rat model | Organoid | hEST |
|---|---|---|---|---|
| ESR1 | 47 | -0.87 | 0 | 0 |
| FGF8 | 43 | 0 | 0 | 1 and 7 |
| KRAS | 41 | -0.17 | 0.5852106 | 0 |
| TNF | 41 | 0 | 0 | 7 |
| CCND1 | 40 | 0 | 0.3452067 | 0 |
| CASP3 | 38 | 0 | -0.4967571 | 7 |
| TGFB1 | 38 | 0 | 1.0573403 | 0 |
| SMAD3 | 37 | 0 | -0.4921032 | 0 |
| EP300 | 36 | 0 | 0 | 7 |
| CDH1 | 35 | 0 | 1.1117428 | 0 |
| GLI1 | 32 | 0 | -0.5652563 | 0 |
| FGF18 | 31 | 0 | -1.3078685 | 1 |
| POU5F1 | 30 | 0 | 0.8828229 | 0 |
| FGF3 | 29 | 0 | 1.1822592 | 0 |
| FGF20 | 28 | 0 | **0** | 0 |
| ABL1 | 24 | 0 | -0.3836435 | 1 and 7 |
| AGT | 24 | 0 | **0** | 0 |
| PTCH1 | 24 | 0 | 0.3741653 | 7 |
| SMO | 23 | 0.21 | -0.3740355 | 1 |
| IGF2 | 22 | 0 | -0.6497496 | 0 |
| RAD51 | 22 | 0.26 | 0.3817702 | 0 |
| NOTCH2 | 21 | 0 | 0 | 0 |
| PLK1 | 21 | 0.35 | 0.3320304 | 0 |
| TGFBR1 | 21 | 0.16 | -0.6739545 | 0 |
| HAND2 | 20 | 0 | 0 | 0 |

**C**

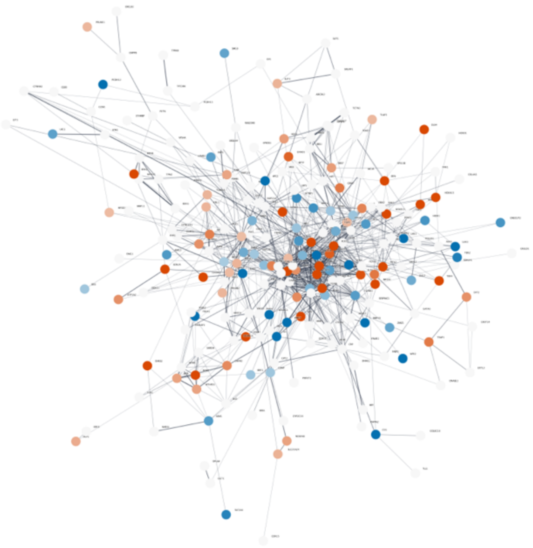

**D**

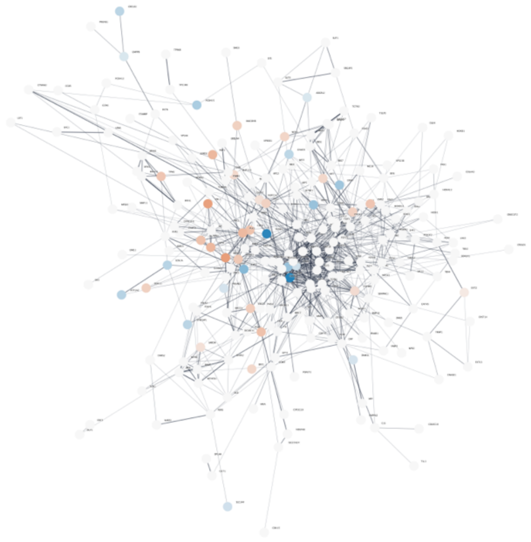

**Supplementary Figure S4 (A)** The BD-hub genes are coloured based on evidence of differential expression in any of 3 published studies (red) or not (grey). **(B)** Table of most connected genes, ranked by degree (number of direct neighbours in the BD-hub) in column 2, columns 3 and 4, are logFC (red for upregulated, blue for downregulated) for the rat and organoid exposure models, and column 4 shows dysregulation at day 1,7 or both timepoints as reported in the embryonic stem cell toxicity test. **(C)** Genes dysregulated in human forebrain organoid of autism risk, orange=upregulated, blue=downregulated, strength of colour = size of logFC. **(D)** Genes dysregulated in the Rat model of VPA-induced neurodevelopmental disability, orange=upregulated, blue=downregulated, strength of colour = size of logFC.

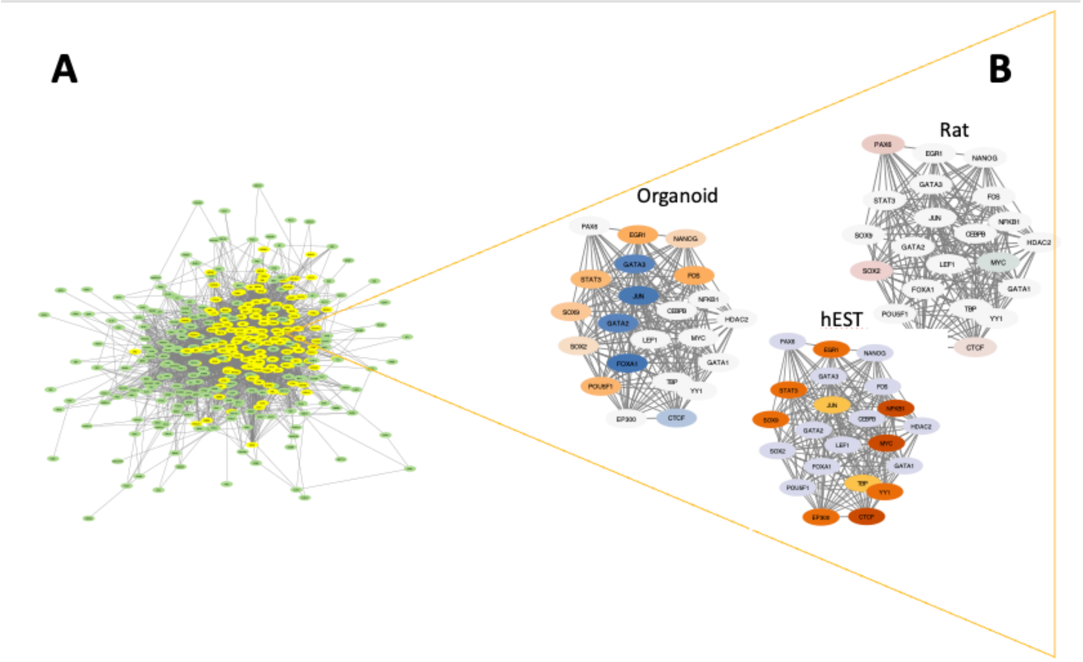


**Supplementary Figure S5: (A)** Protein-protein interaction (PPI) network derived from TFs with binding affinity that is predicted to be modified by genetic variation within the VPA-exposed BD-hub genes. The most highly connected genes (N=22, degree range = 71 -147), considered to be master regulators, are highlighted in yellow. **(B)** Overlay on of differential expression, on the 22 highly connected genes, from human forebrain organoid of autism risk (Organoid) and rat model of VPA-induced neurodevelopmental disability (Rat), orange=upregulated, blue=downregulated, strength of colour = size of logFC and the embryonic stem cell toxicity test (hEST), data at day 1 (orange), 7 (dark orange) or both timepoints (brown).

**Exome capture at high confidence variant loci**

Comparison of the mean read depths (Wilcoxon signed-rank test, Supplementary Fig. 6) at the loci of high-confidence variants in VPA-exposed samples found no significant difference between Nextera and the Broad institute custom kit (*n*= 51 vs *n* =12 [95% CI -5.00 to 7.00], *p* = 0.85) or NimbleGen (*n* = 3, [95% CI -11.0 to 22.0], *p* = 0.865) or between NimbleGen and the Broad custom kit ([95% CI -9.99 to 13.99], *p* = 0.51).

**A**

| | NimbleGen | Nextera | Broad_custom | Total |
|---|---|---|---|---|
| **Cases** | 3 | 24 | 1 | 28 |
| **Controls** | 0 | 27 | 11 | 38 |
| **Total** | 3 | 51 | 12 | 66 |

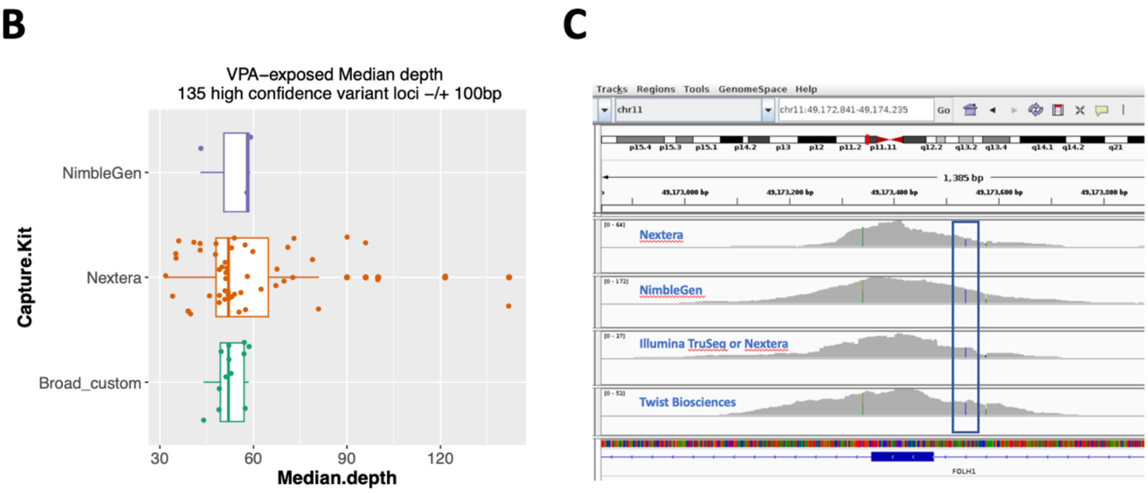


**Supplementary Fig. S6 (A)** Table of Capture kits used for exome sequencing. **(B)** box plots of median read coverage at high-confidence variant loci from samples of pregnancies exposed to VPA. **(C)** Screenshot of IGV browser showing bam files for 4 samples for which different exome capture kits had been applied. The blue rectangle highlights the region where the *FOLH1* intronic variant was identified.

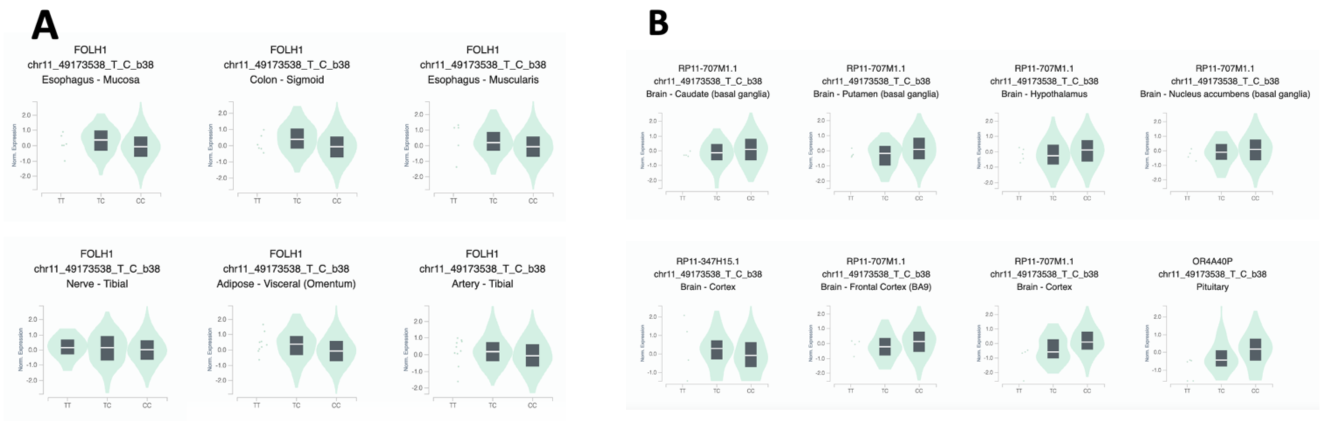


**Supplementary Figure S7:** eQTL violin plots, showing expression differences between alleles for variant chr11:49173538:T:C observed in different tissues for *FOLH1* **(A)** and other transcripts **(B)** as detailed below:

| Gene Symbol | P-Value | NES | Tissue |
|---|---|---|---|
| RP11-347H15.1* | 0.00002 | -0.42 | Brain - Cortex |
| FOLH1 | 2.60E-07 | -0.4 | Colon - Sigmoid |
| FOLH1 | 3.20E-08 | -0.39 | Esophagus - Mucosa |
| FOLH1 | 0.000084 | -0.27 | Esophagus - Muscularis |

| | | | |
|---|---|---|---|
| FOLH1 | 0.00057 | -0.23 | Adipose - Visceral (Omentum) |
| FOLH1 | 0.00096 | -0.21 | Artery - Tibial |
| FOLH1 | 0.000084 | -0.12 | Nerve - Tibial |
| RP11-707M1.1* | 0.000017 | 0.35 | Brain - Nucleus accumbens (basal ganglia) |
| RP11-707M1.1 | 0.0000016 | 0.36 | Brain - Putamen (basal ganglia) |
| OR4A40P | 0.000066 | 0.38 | Pituitary |
| RP11-707M1.1 | 9.80E-08 | 0.42 | Brain - Caudate (basal ganglia) |
| RP11-707M1.1 | 0.000028 | 0.42 | Brain - Frontal Cortex (BA9) |
| RP11-707M1.1 | 0.0000077 | 0.43 | Brain - Hypothalamus |
| RP11-707M1.1 | 0.000037 | 0.47 | Brain - Cortex |

* Transcribed_unprocessed_pseudogene: Long non-coding RNA transcripts that do not encode a functional protein, but may have other roles in the cell

#### *EP300* coexpression: development of a reference map

**A.**

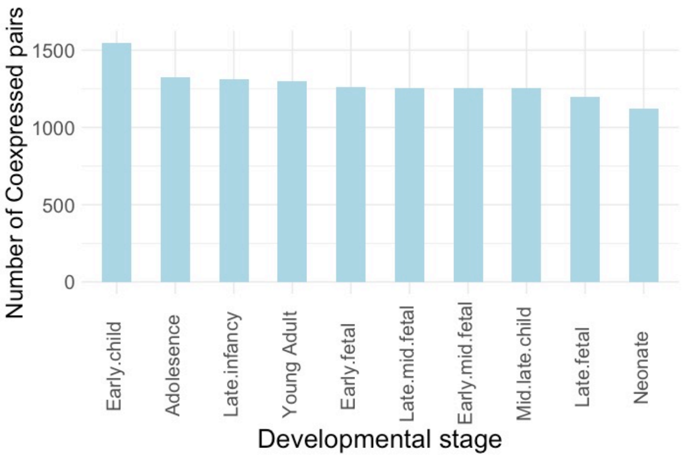


**B.**

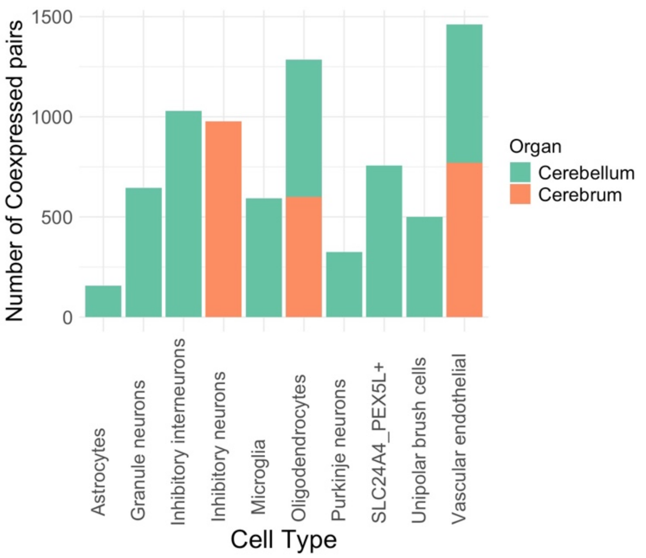


**Supplementary Figure S8: Profiling EP300-TF coexpression. (A)** Histogram showing the number of *EP300*-TF pairs with correlated expression (P < 0.05 for Pearson or Spearman correlation tests) based on RNA-seq data representing brain tissues across developmental stages obtained from the BrainSpan (https://www.brainspan.org/) resource. (B) The number of *EP300*-TF pairs with correlated expression observed in modules identified using the high definition Weighted Gene Coexpression Network Analysis (hdWGCNA) method and scRNA-seq data representing foetal brain cell types.

### RNA-seq analysis in hESC-derived neurons

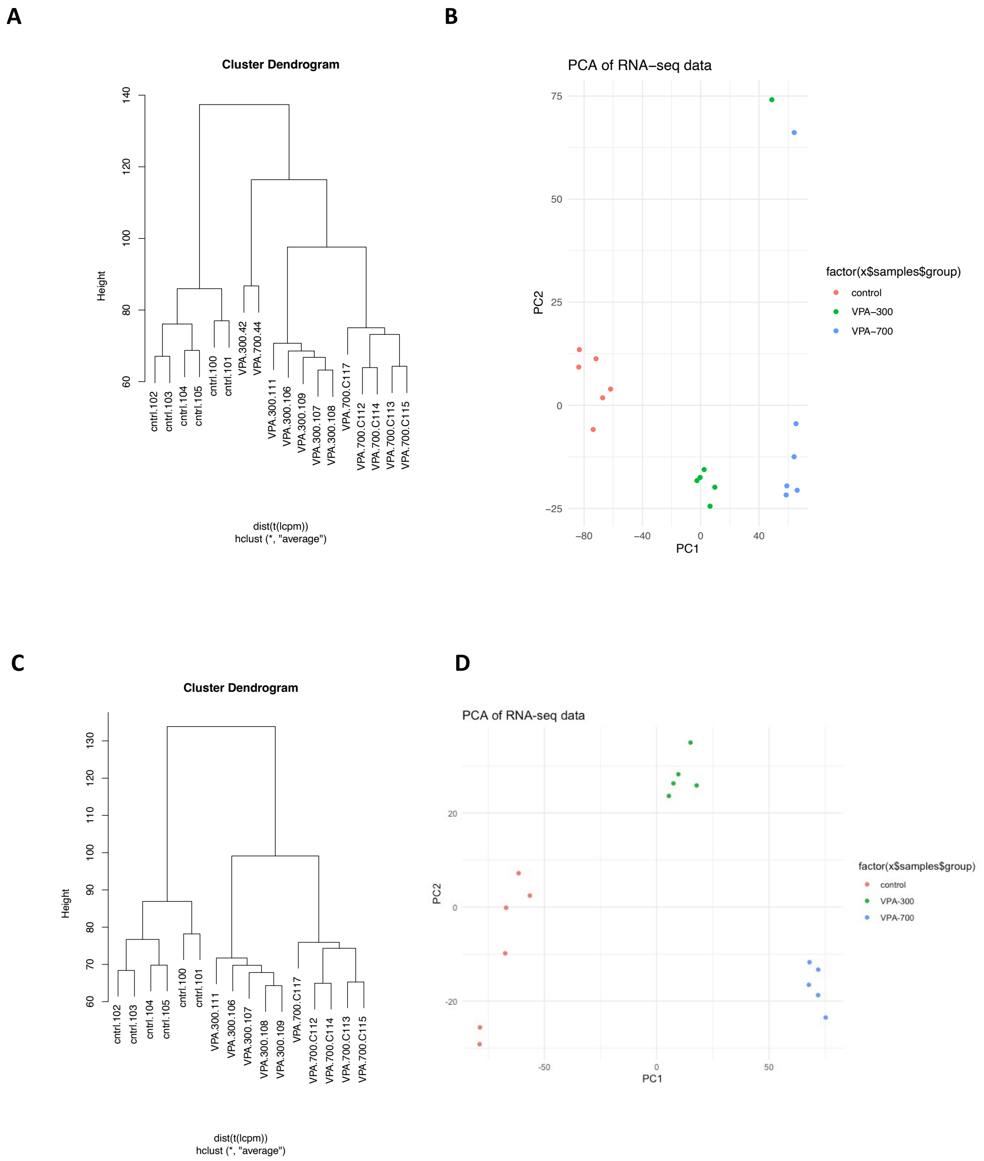


**Supplementary Figure S9:** The hierarchical cluster analysis (**A**) and PCA plot (**B**) for all 18 samples showed two outliers that were removed from further analysis. The hierarchical cluster diagram (**C**) and PCA plot (**D**) for the remaining 16 samples showed clear clustering of comparison groups.

### Differential expression analysis in hESC-derived neurons

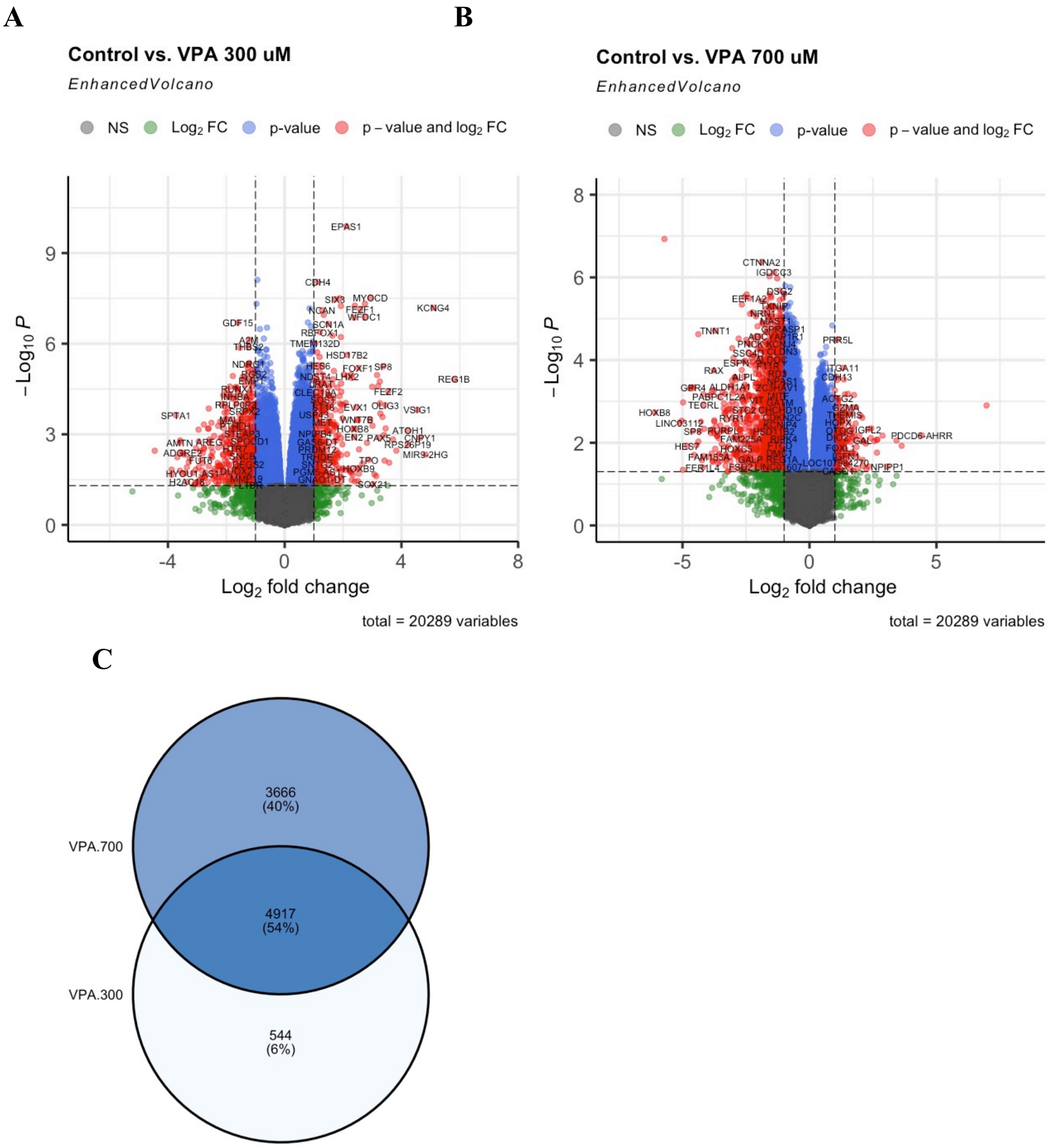


**Supplementary Figure S10: Results from differential expression analyses. (A)** Control vs. VPA 300 µM. (**B**) Control vs. VPA 700 µM. (**C**) Venn diagram showing overlap in dysregulated genes at different doses (top circle = VPA 700 µM, bottom circle = VPA 300 µM,).

### References


1. Rappaport N, Twik M, Plaschkes I, et al. MalaCards: an amalgamated human disease compendium with diverse clinical and genetic annotation and structured search. Nucleic Acids Res. 2017;45(D1):D877-D87.
2. Consortium EP. An integrated encyclopedia of DNA elements in the human genome. Nature. 2012;489(7414):57-74.
3. Feleke R, Jazayeri D, Abouzeid M, et al. Integrative genomics reveals pathogenic mediator of valproate-induced neurodevelopmental disability. Brain. 2022;145(11):3832-42.

4. Cui K, Wang Y, Zhu Y, et al. Neurodevelopmental impairment induced by prenatal valproic acid exposure shown with the human cortical organoid-on-a-chip model. Microsystems & nanoengineering. 2020;6(1):1-14.
5. Meng Q, Zhang W, Wang X, et al. Human forebrain organoids reveal connections between valproic acid exposure and autism risk. Translational psychiatry. 2022;12(1):1-8.
6. Schulpen SHW, Pennings JLA, Piersma AH. Gene Expression Regulation and Pathway Analysis After Valproic Acid and Carbamazepine Exposure in a Human Embryonic Stem Cell-Based Neurodevelopmental Toxicity Assay. Toxicol Sci. 2015;146(2):311-20.
